# Plasma proteome-wide analysis of cerebral small vessel disease identifies novel biomarkers and disease pathways

**DOI:** 10.1101/2024.10.07.24314972

**Authors:** Gabriela T. Gomez, Liu Shi, Alison E. Fohner, Jingsha Chen, Yunju Yang, Myriam Fornage, Michael R. Duggan, Zhongsheng Peng, Gulzar N. Daya, Adrienne Tin, Pascal Schlosser, W.T. Longstreth, Rizwan Kalani, Malveeka Sharma, Bruce M. Psaty, Alejo J. Nevado-Holgado, Noel J. Buckley, Rebecca F. Gottesman, Pamela L. Lutsey, Clifford R. Jack, Kevin J. Sullivan, Thomas Mosley, Timothy M. Hughes, Josef Coresh, Keenan A. Walker

**Author notes:** **Corresponding Author:**Keenan A. Walker, 251 Bayview Boulevard, Room 04B316, Baltimore, MD 21224.

## Abstract

Cerebral small vessel disease (SVD), as defined by neuroimaging characteristics such as white matter hyperintensities (WMHs), cerebral microhemorrhages (CMHs), and lacunar infarcts, is highly prevalent and has been associated with dementia risk and other clinical sequelae. Although conditions such as hypertension are known to contribute to SVD, little is known about the diverse set of subclinical biological processes and molecular mediators that may also influence the development and progression of SVD. To better understand the mechanisms underlying SVD and to identify novel SVD biomarkers, we used a large-scale proteomic platform to relate 4,877 plasma proteins to MRI-defined SVD characteristics within 1,508 participants of the Atherosclerosis Risk in Communities (ARIC) Study cohort. Our proteome-wide analysis of older adults (mean age: 76) identified 13 WMH-associated plasma proteins involved in synaptic function, endothelial integrity, and angiogenesis, two of which remained associated with late-life WMH volume when measured nearly 20 years earlier, during midlife. We replicated the relationship between 9 candidate proteins and WMH volume in one or more external cohorts; we found that 11 of the 13 proteins were associated with risk for future dementia; and we leveraged publicly available proteomic data from brain tissue to demonstrate that a subset of WMH-associated proteins was differentially expressed in the context of cerebral atherosclerosis, pathologically-defined Alzheimer’s disease, and cognitive decline. Bidirectional two-sample Mendelian randomization analyses examined the causal relationships between candidate proteins and WMH volume, while pathway and network analyses identified discrete biological processes (lipid/cholesterol metabolism, NF-kB signaling, hemostasis) associated with distinct forms of SVD. Finally, we synthesized these findings to identify two plasma proteins, oligodendrocyte myelin glycoprotein (OMG) and neuronal pentraxin receptor (NPTXR), as top candidate biomarkers for elevated WMH volume and its clinical manifestations.

---

Cerebral small vessel disease (SVD) is a major cause of ischemic stroke and a significant contributor to cognitive decline and dementia.^1–3^ Consequences of SVD can be characterized using neuroimaging as white matter hyperintensities (WMHs), cerebral microhemorrhages (CMHs), and lacunar infarcts.^4,5^ WMHs,^6–8^ CMHs,^9^ and lacunar infarcts^10^ have been associated with poor cognitive outcomes and dementia risk, and both the presence and progression of SVD lesions have been linked to decline in cognitive function.^11^ SVD is a central feature of vascular cognitive impairment and dementia (VCID),^12^ and certain forms of SVD have been shown to increase risk for AD dementia as well.^13^ Notably, WMHs, appearing decades before onset of clinical symptoms, not only confer increased risk for Alzheimer’s disease (AD), but may also constitute an integral feature of AD pathogenesis.^14^ Beyond dementia and cognitive impairment,^15^ SVD has also been associated with physical/motor decline^16^ and affective dysfunction,^17^ underscoring the clinical relevance of these brain changes.

Understanding the complex molecular mechanisms underlying SVD and its capacity to engender neurodegenerative and clinical sequelae is critical. It is clear that peripheral processes, including vascular disease/risk factors (e.g., hypertension, smoking) and cardiometabolic dysfunction (e.g., obesity, diabetes) confer increased risk for SVD and SVD progression.^11^ However, much of the explanatory variance in SVD risk is still unaccounted for. This is underscored by the finding that generally healthy individuals can display SVD, even during early and middle adulthood.^18^ Genome-wide association studies (GWAS’s) have identified susceptibility loci for SVD and stroke. These studies support the etiological relevance of immune and inflammatory pathways,^19,20^ cell signaling and cell survival,^21^ and endothelial function.^22^ Given the relevance of systemic factors to cerebrovascular health, a data-driven analysis of the plasma proteome may facilitate a deeper understanding of peripheral biological changes that promote SVD and may highlight novel plasma biomarkers that can non-invasively identify individuals at risk for VCID.

Though the roles of discrete proteins have been investigated in the context of SVD, the plasma proteomic signature of SVD burden is largely unexplored. The current study utilized SomaScan Multiplexed Proteomic technology^23,24^ to examine the relationship between the plasma level of nearly 5000 proteins and MRI-defined SVD within the Atherosclerosis Risk in Communities (ARIC) Study Cohort. This proteome-wide association study identified 13 proteins associated with WMH volume and numerous candidate proteins showing suggestive associations with lacunar infarcts and cerebral microhemorrhages. After reproducing protein-WMH associations using multiple external cohorts, we incorporated systems level analyses to identify biologically relevant protein networks and understand the peripheral biological processes and regulatory mechanisms underlying SVD pathogenesis. Lastly, we used two-sample Mendelian randomization to identify the causal relationships between candidate plasma proteins, protein networks, and WMH volume.

## Results

### Sample characteristics

A total of 1508 ARIC participants were included in the primary analytic sample (age: 76.3 years [SD: 5.22]; 58.6% women; 25.1% Black; **Figure 1**; **Supplementary Table 1**). At the time of MRI and protein measurement (ARIC visit 5; 2011-2013) the prevalence of lacunar infarcts was 18.2%, and subcortical CMHs and lobar CMHs were present in 19.2% and 8.9% of participants, respectively; the mean WMH volume was 9.37mm^3^. A study flowchart with participant inclusion/exclusion criteria is outlined in **Extended Data Figure 1**. Of the 5,284 available aptamer binding reagents, 4,877 (91%) passed quality control (refer to Methods) and were included in the current analysis.

**Figure 1.**
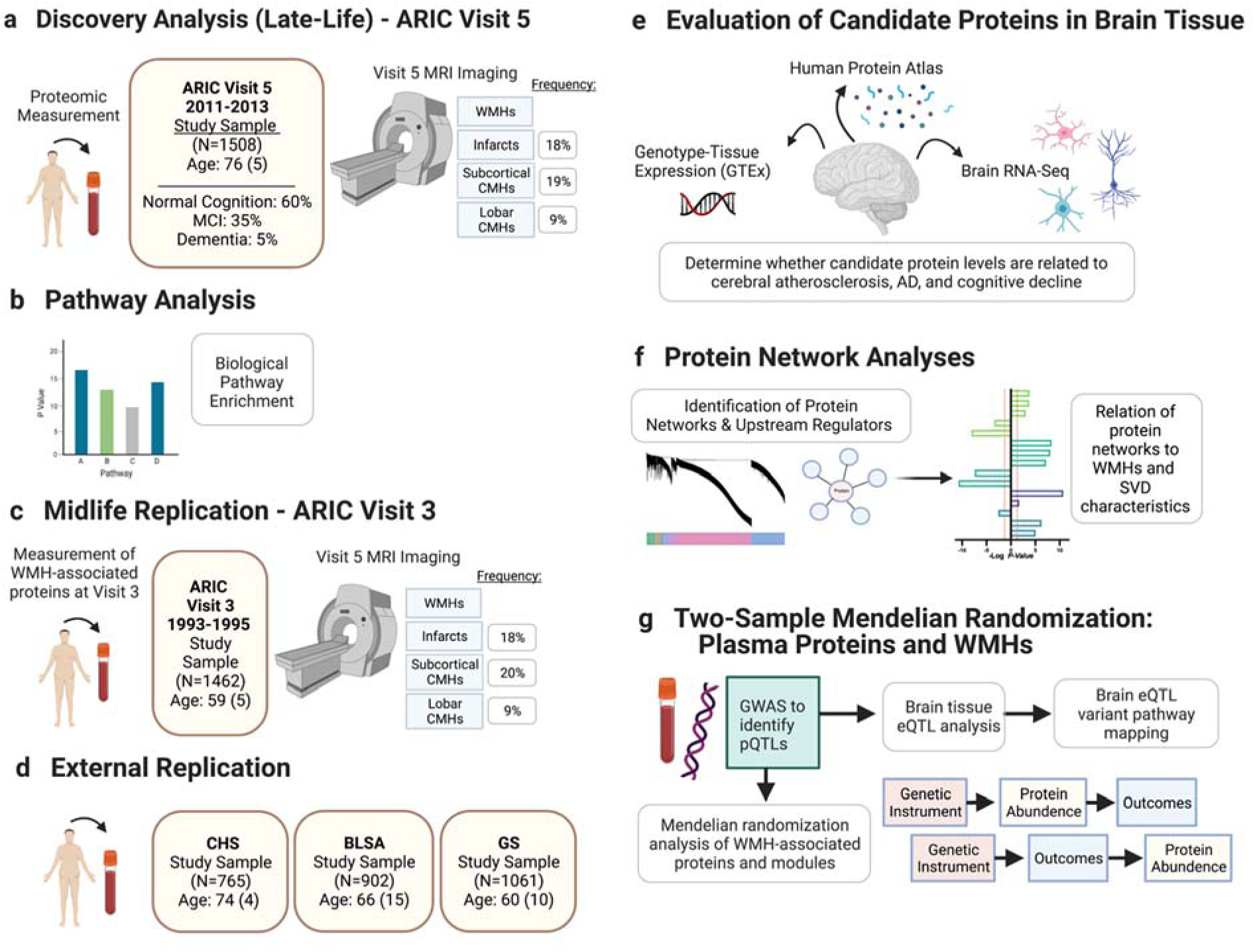
Schematic overview of study design. **a,** The primary analysis related proteins measured at ARIC visit 5 (late-life baseline; 2011-2013) to neuroimaging characteristics measured at visit 5. **b,** Ingenuity Pathway Analysis was used to identify enriched biological pathways and protein networks among the top WMH-associated proteins. **c,** Midlife replication analyses related the midlife (ARIC visit 3; 1993-1995) levels of the SVD-associated proteins (identified in the primary analysis) to WMH volume measured approximately two decades later at ARIC visit 5. **d,** WMH-associated proteins were replicated in external cohorts. **e,** Genotype-Tissue Expression (GTEx), Human Protein Atlas, and Brain RNA-Seq databases were used to identify SVD-associated proteins expressed in brain tissue and determine cellular specificity. SVD-associated proteins were examined for their relationship with cerebral atherosclerosis, pathologically-defined Alzheimer’s disease, and cognitive decline using computed results from available brain proteomic studies. **f,** Networks of co-expressed proteins were identified using the data-driven Netboost clustering method and then related to small vessel disease neuroimaging characteristics. **g,** A genome-wide association study was conducted to identify loci associated with plasma levels of the top SVD-associated proteins. Mendelian randomization analyses incorporating pQTLs were used to explore causal associations with WMH volume. *Abbreviations:* ARIC, Atherosclerosis Risk in Communities Study; eQTLs, expression quantitative trait loci; pQTLs, protein quantitative trait loci; SVD, small vessel disease; WMH, white matter hyperintensity

### Proteome-wide analysis identifies multiple WMH-associated proteins

We first examined the association between levels of the 4,877 proteins measured during late-life (visit 5) and concurrently measured WMH volume. In unadjusted linear regression analyses, 442 proteins were associated with WMH volume at an FDR-corrected *P*-value <0.05. After adjustment for demographic variables, kidney function, and cardiovascular risk factors, 13 proteins remained significantly associated with WMH volume at an FDR-corrected threshold (**Figure 2A; Supplementary Table 2**). Oligodendrocyte-myelin glycoprotein (OMG), a protein implicated in cell adhesion, neuronal regeneration and axonogenesis, exhibited the strongest association – by *p*-value – with WMH volume (*ß*= –0.167, *p*=4.57×10^−7^). Neuronal Pentraxin Receptor (NPTXR), a regulator of postsynaptic neurotransmitter receptor activity, also demonstrated a strong relationship with WMH volume (*ß*= –0.247, *p*=9.80×10^−7^). Lower levels of both OMG and NPTXR were associated with increased WMH volume. Among the other top WMH-associated proteins, several function in cell proliferation and apoptosis/autophagy (ESM1, HSPA1B, TNFRSF11B), signal transduction (EPHA10, RSPO1), cell differentiation and organ development (CHRDL1, MPST, REN, RSPO1, TAGLN), and synaptic regulation (CLSTN3). See **Supplementary Table 3** for more detailed biological processes, molecular functions, and disease associations of each top WMH-associated protein. In a sensitivity analysis additionally adjusting for *APOE* ε4 status, all but one protein (RSPO) remained statistically significant at the same FDR-corrected threshold (**Supplementary Table 4)**. Results remained similar after adjusting for prevalent stroke (**Figure 2B**) and after incorporating sampling weights to account for selection for ARIC visit 5 MRI (**Supplementary Table 4**). Given that some cardiovascular risk factors may serve as mediators in the protein-WMH relationship, we additionally examined models without cardiovascular risk factor adjustment. These results implicate a broader group of statistically significant proteins, though OMG and NPTXR remained the top two candidate proteins (**Supplementary Table 5)**. Notably, we found that genetic variants linked to 7 of the 13 identified WMH-associated proteins were also associated with one or more cardiovascular or metabolic traits, including blood pressure and cholesterol level (**Figure 2C**).

**Figure 2.**
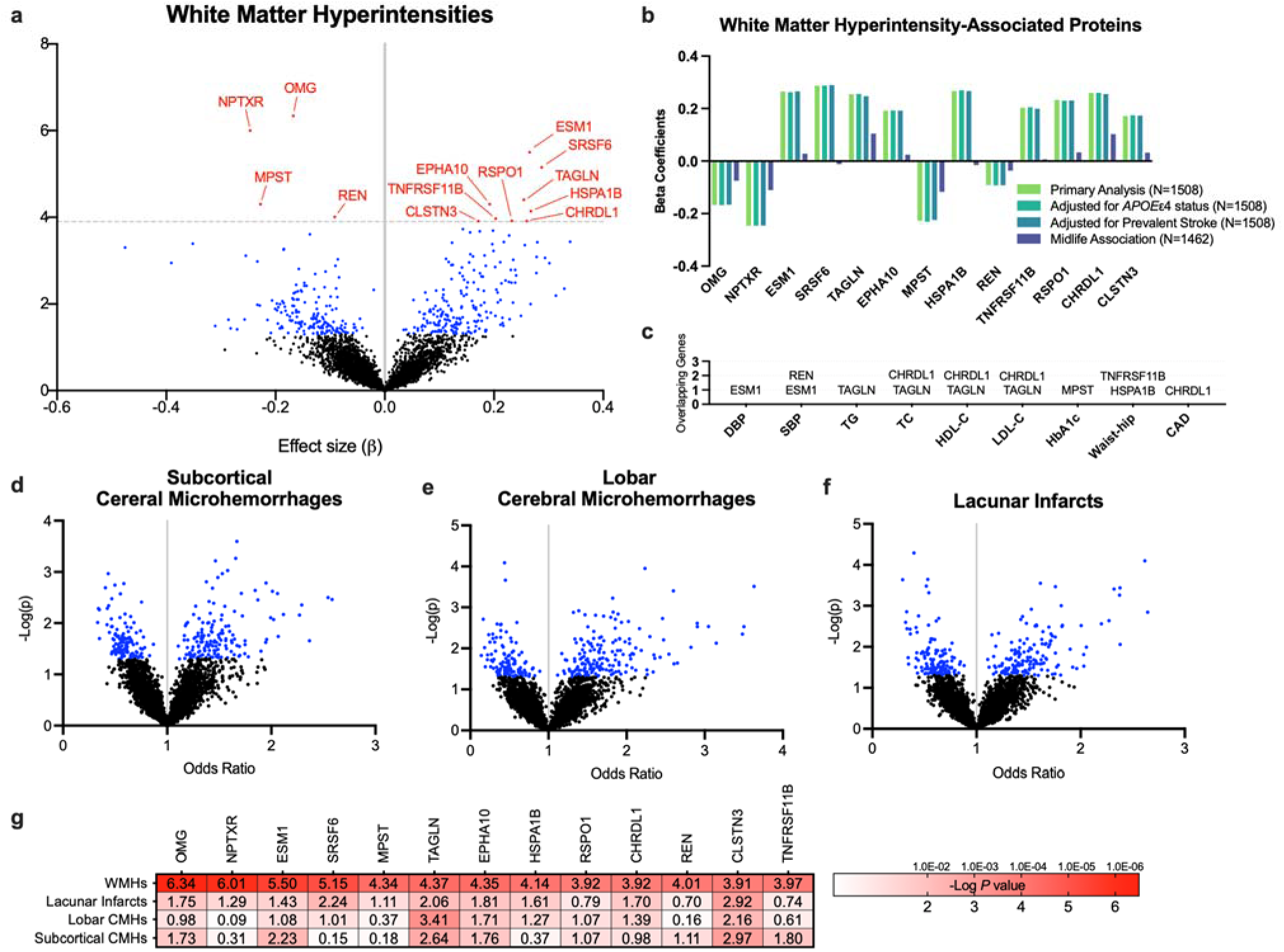
Proteome-wide associations with MRI-defined cerebral small vessel disease. A proteomic analysis of cerebral small disease characteristics was performed using multivariable linear and logistic regression models adjusted for age, sex, race-center, education, estimated glomerular filtration rate (eGFR)-creatinine, body mass index, diabetes, hypertension, and smoking status at the time of protein assessment. Volcano plots display the effect sizes (x-axis) and p-values (y axis) for the association of log2 protein level with **a,** white matter hyperintensity (WMH) volume and the presence of **d,** subcortical cerebral microhemorrhages (CMHs), **e,** lobar CMHs, and **f**, lacunar infarcts. Proteins in red were significant at the FDR-corrected *P*<0.05 threshold; proteins in blue were significant at an uncorrected *P*<0.05 threshold. **b,** After adjusting for prevalent stroke, all WMH-associated proteins remained significant at an uncorrected *p*<0.05 threshold. When the 13 WMH-associated proteins were measured at midlife, two proteins (OMG and NPTXR) remained associated with late-life WMH volume at an uncorrected *p*<0.05 threshold. **c,** Proteins significantly associated with WMH volume and coded for by genes associated in GWAS with cardiovascular and metabolic traits and disease. Gene lists were based on GWAS catalog summary statistics.^78^ **g,** Among the 13 WMH-associated proteins, 8 were associated with lacunar infarcts, 6 with subcortical CMHs, and 4 with lobar CMHs at an uncorrected threshold of *P*<0.05. *Abbreviations:* CAD, coronary artery disease; DBP, diastolic blood pressure; HbA1c, hemoglobin A1C; HDL-C, high-density lipoprotein cholesterol; LDL-C, low-density lipoprotein cholesterol; SBP, systolic blood pressure; TC, total cholesterol; TG, triglycerides; Waist-hip, waist-to-hip ratio

We also conducted a proteome-wide analysis of other SVD phenotypes: lacunar infarcts, subcortical CMHs, and lobar CMHs, each of which was characterized as present (one or more) vs. absent on brain MRI. Although a number of proteins were associated with lacunar infarcts and lobar CMHs in unadjusted analyses, these associations did not remain statistically significant at the proteome-wide FDR-corrected threshold after adjusting for demographic variables, kidney function, and cardiovascular risk factors (**Figure 2D-2F**). The top proteins (at an uncorrected *P*<0.01) and their associations with lacunar infarcts, subcortical CMHs, and lobar CMHs are provided in **Supplementary Table 6**. Based on the hypothesis that distinct forms of SVD have a shared etiology,^25^ we next examined whether levels of the 13 WMH-associated proteins were also associated with the presence of lacunar infarct or CMHs. As shown in **Figure 2G**, 8 of the 13 WMH-associated proteins were associated with lacunar infarcts, 6 with subcortical CMHs, and 4 with lobar CMHs at an uncorrected threshold of *P*<0.05. Each of these associations was directionally consistent with the protein-WMH associations.

### Cerebral small vessel disease subtypes implicate distinct biological pathways

To further enhance our understanding of the biological pathways associated with each MRI-defined measure of SVD, we conducted a series of pathway analyses using SVD-associated proteins. Proteins associated with each SVD outcome at an unadjusted *P*<0.01 (**Supplementary Table 6**) were examined for canonical (biological) pathway enrichment using the Ingenuity Pathway Analysis (IPA) tool. WMH-associated proteins were enriched for LXR/RXR signaling (involved in cholesterol metabolism), coagulation, atherosclerosis signaling, and activation of the SPINK1 pancreatic cancer pathway (**Figure 3**). Proteins associated with lobar CMH were similarly enriched for coagulation and extrinsic/intrinsic prothrombin pathways, whereas proteins associated with subcortical CMH demonstrated enrichment for a distinct set of biological pathways, including cardiac hypertrophy and PI3K/AKT signaling. On the other hand, proteins associated with lacunar infarcts were enriched for immune/inflammatory pathways, specifically NF-kB and acute phase response signaling. A broader set of biological pathways was enriched among SVD-associated proteins in analyses that used the full gene set in the IPA database as the reference (see **Extended Data Fig. 2**).

**Figure 3.**
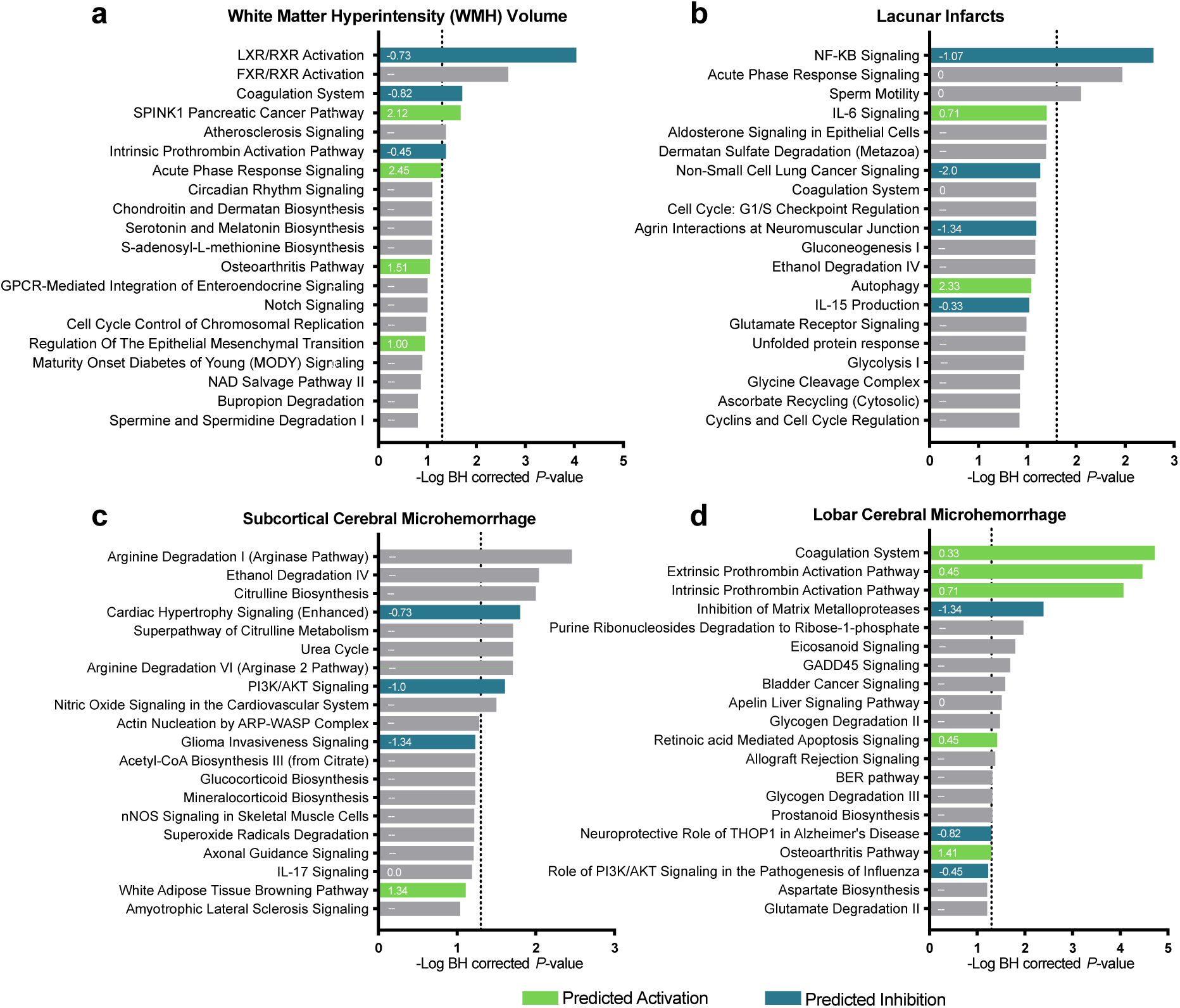
Enriched biological pathways for proteins associated with cerebral small vessel disease. For each small vessel disease characteristic—white matter hyperintensities, lacunar infarcts, subcortical and lobar cerebral microhemorrhages—the top 25 canonical pathways, identified using Ingenuity Pathway Analysis (IPA), are displayed. The threshold for statistical significance, defined as an FDR-corrected *P*<0.05, is represented by the vertical dotted line. The Z-score indicating the predicted degree of activation (green) or inhibition (blue) for each pathway is displayed within each bar. For bars with no Z-scores, the extent of activation could not be predicted. These results incorporate the full set of SomaScan proteins included in the study as the reference gene set.

### Midlife levels of OMG and NPTXR are associated with late-life WMH volume

Next, we examined whether the 13 WMH-associated proteins identified in our primary analysis maintained an association with late-life WMH volume when measured 18 years prior during middle adulthood (ARIC visit 3; 1993-1995; mean age: 58.7 [SD: 5.2]). A total of 1462 participants who received a 3T brain MRI during late-life (ARIC visit 5) also had SomaScan proteomic measurement during midlife (see **Extended Data Fig. 1**; participant characteristics in **Supplementary Table 1**). Of the 13 WMH-associated proteins examined, OMG and NPTXR were associated with future WMH volume when measured at midlife, after adjusting for demographic variables and cardiovascular risk factors (**Figure 2B; Supplementary Table 2**). This directionally consistent, albeit comparatively weaker, association with late-life WMH volume suggests OMG and NPTXR may serve as prognostic indicators of late-life SVD, even when measured during middle adulthood.

### WMH-associated proteins are replicated in external cohorts

We examined the reproducibility and generalizability of protein-WMH associations in three external cohorts. Using data from the Cardiovascular Health Study (CHS), a cohort of older adults demographically similar to the ARIC cohort with SomaScan v.4 protein measurements and 1.5T brain MRI scans (n=765; mean age: 73.6 [SD 4.4]), we found that 8 of the 13 candidate proteins were associated with WMH volume using a threshold of *P*<0.05 (uncorrected) after adjusting for potential confounders. Using a partially overlapping sample of CHS participants with serial MRI scans (n=680), we found that 5 of the 13 candidate proteins were associated with worsening white matter grade, an index of progressive white matter lesions, over a mean 5-year follow-up (SD: 0.6 years) period (**Supplementary Tables 7-8**). In total, 11 out of 13 proteins were associated with at least one WM outcome in CHS; these results were directionally consistent with that of the discovery analysis (**Extended Data Fig. 3**).

Using data from the Baltimore Longitudinal Study of Aging (BLSA) – a younger and healthier cohort with SomaScan v.4.1 protein measurements and 3T MRI data (n=902; mean age: 66.1 [SD 14.7]) – we found that 3 of the 13 candidate proteins (OMG, NPTXR, TNFRSF11B) were significantly associated with WMH volume after adjusting for potential confounders (**Supplementary Tables 9-10**). Of the 13 candidate proteins another 3 (TAGLN, RSPO1, CHRDL1) were significantly associated with total white matter volume, a distinct, yet related, metric. These findings were directionally consistent with the discovery results (**Extended Data Fig. 3**). In a cohort of cognitively normal middle-aged adults with SomaScan v.4 protein measurements and 3T MRI data from the Generation Scotland study (n=940 mean age: 59.9 [SD 9.6]), we found that 2 of the 10 available candidate proteins (HSPA1B and TNFRSF11B) were associated with total WMH volume in a manner directionally consistent with the discovery results (**Supplementary Tables 11-12**). In total, 9 protein-WMH associations replicated in at least 1 external cohort. Three proteins replicated in at least 2 external cohorts: OMG, NPTXR, and TNFRSF11B.

### Select WMH-associated proteins are expressed in brain tissue

To determine the likely origin of WMH-associated proteins, we next examined RNA and protein expression across tissues using the GTEX database and data from the Human Protein Atlas, respectively. As illustrated in **Figure 4A and 4B**, top WMH-associated proteins, OMG and NPTXR, are selectively, or at least preferentially, detected in brain tissue, suggesting that the OMG and NPTXR proteins measured in plasma likely largely originate from the CNS. TNFRSF11B protein levels, on the other hand, were not detected at meaningful levels in brain tissue. We did find, however, that TNFRSF11B is expressed at an RNA level across numerous tissue types, especially arterial tissue, suggesting a role for this transcript in vascular processes. Using publicly available cell-specific human brain RNA-seq data (Brain RNA-Seq)^26^, we identified *OMG* as preferentially expressed by oligodendrocytes, whereas *NPTXR* was preferentially expressed by neurons (**Figure 4C**).

**Figure 4.**
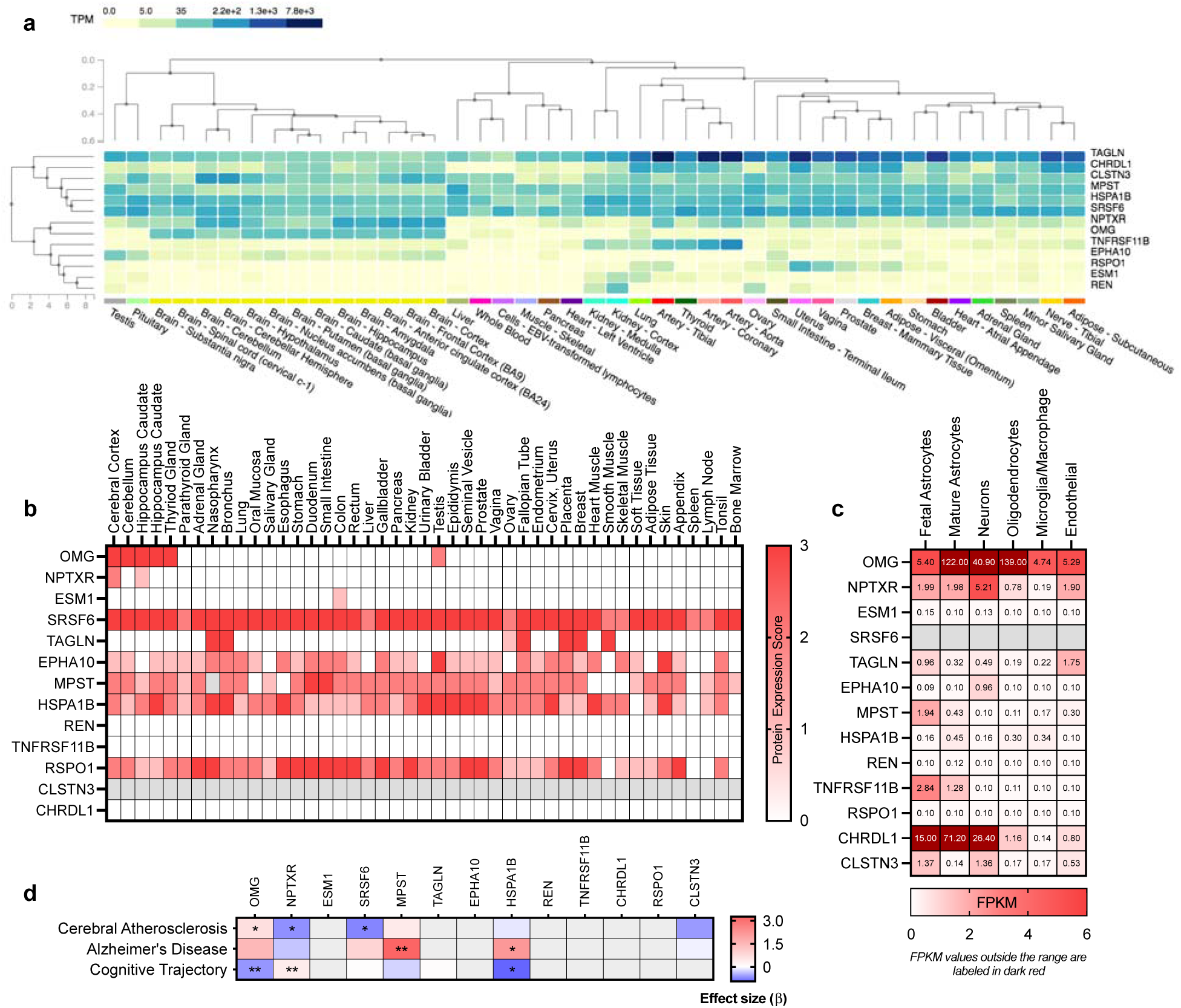
WMH-associated proteins are expressed in brain and associated with cerebral atherosclerosis. **a,** Gene expression in brain, whole blood, and other tissues using data available from postmortem samples in the GTEX database^79^. Gene expression is reported in transcripts per million. Genes and tissues are displayed according to hierarchical clustering. **b,** Protein expression in brain and other tissue using data available in the Tissue Atlas of the Human Protein Atlas.^80^ The Antibody-based protein levels are qualitatively displayed as (0) not detected, (1) low, (2) medium, and (3) high. https://www.proteinatlas.org/ **c,** Brain cell-specific expression of genes coding for WMH-associated proteins in humans derived from the Brain RNA-Seq database.^81^ Values are reported in fragments per kilobase per mission reads (FKPM). http://www.brainrnaseq.org/ **d,** Results from external brain proteomic studies for candidate proteins in brain tissue. Results derived from Wingo et al. 2019^28^ (cerebral atherosclerosis) and Wingo et al. 2020^27^ (Alzheimer’s disease and cognitive trajectory). Heatmap displays effect sizes (beta). \**P*<0.05; \*\**P*<0.01.

### Brain levels of WMH-associated proteins are associated with disease phenotypes

As several WMH-associated proteins are highly expressed in brain tissue, we utilized published results from the Religious Orders Study and the Rush Memory and Aging Project (ROS/MAP), BLSA, and Banner cohorts^27,28^ to understand how the expression of these proteins in brain relates to cerebral atherosclerosis, Alzheimer’s disease, and cognitive decline. Of the 6 WMH-associated proteins measured by ROS/MAP in brain tissue, 3 were significantly associated with cerebral atherosclerosis after adjusting for other neuropathologies (**Figure 4D**; **Supplementary Table 13**). Brain OMG levels were positively associated with cerebral atherosclerosis, whereas brain NPTXR and SRSF6 levels were lower among participants with elevated cerebral atherosclerosis. Within the same cohort, brain levels of WMH-associated proteins OMG, NPTXR, and HSPA1B were associated with cognitive decline, whereas MPST and HSPA1B were found to be elevated in pathologically-defined AD brains (versus control brains). Interpreting our findings in the context of these external proteomic analyses lends additional support for the pathological and clinical relevance of the top WMH-associated candidate proteins.

### Protein networks are strongly associated with cerebral small vessel disease

Next, we identified networks of co-expressed proteins using the full set of proteins measured within the ARIC cohort (visit 5). We applied the data-driven Netboost clustering method^29^ to group correlated proteins in 15 unique modules containing 27 to 2665 proteins (**Figure 5A-B; Supplementary Table 14**). Using module specific expression values derived for each participant, we found that six modules were associated with WMH volume, three with lacunar infarcts, and two with CMHs, after adjusting for demographic and cardiovascular risk factors (**Figure 5C**). After FDR corrections for multiple comparisons, module 9 (ME9) and module 10 (ME10) maintained a significant association with WMH, whereas module 12 (ME12) remained associated with lacunar infarcts. Functional profiling of protein modules using g:Profiler^30^ characterized ME9 as enriched for pancreatic secretion, protein digestion, ECM degradation, and complement signaling (**Figure 5D**), whereas ME10 was enriched for proteins responsible for spliceosome function, growth hormone signaling, and IL-11 signaling (**Figure 5E**). Three of the top ME10 hub proteins (EPHA10, CLSTN3, HSPA1B) are also among the top 13 WMH-associated proteins from our discovery analysis, and 5 of the 6 ME10 hub proteins (HSPA1B, EPHA10, CLSTN3, PUF60, SUMF2) were found to be differentially expressed at the RNA or protein level in at least one brain region in AD brains (versus control brains) using the AMP-AD consortium dataset (AMP-AD Sage Bionetworks Agora platform). The module associated with lacunar infarcts (ME12) was strongly enriched for proteins involved in complement signaling, coagulation, and other immune processes (**Figure 5F)**. Functional profiling and tissue enrichment analyses for all SVD-associated modules is provided in **Extended Data Fig. 4** and **Extended Data Fig. 5**, respectively.

**Figure 5.**
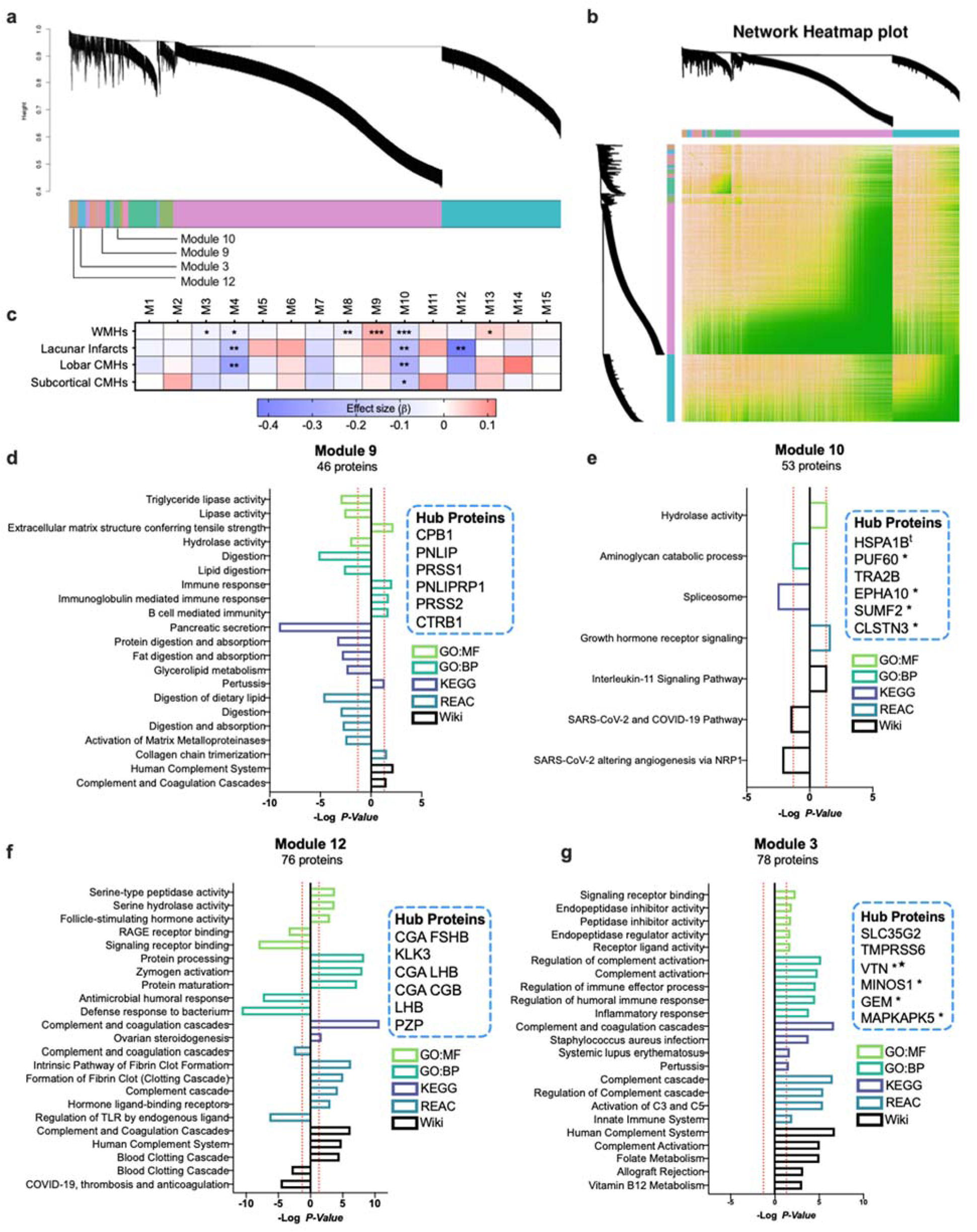
Plasma protein networks are associated with small vessel disease subtypes **a,** Hierarchical cluster tree of 4,877 proteins measured at ARIC visit 5 (2011-13). These proteins are separated into 15 modules using Netboost clustering; this is depicted by the multicolor band. **b,** A heatmap of the Topological Overlap Matrix illustrating the protein networks. Lighter colors represent higher protein-protein adjacency. **c,** A heatmap depicting the association of module expression with cerebral small vessel disease variables; *P*-values: \**P*<0.05; \*\**P*<0.01; \*\*\**P*<0.001. **d-g**, Gene set enrichment (using the g:Profiler toolkit) of each WMH-associated protein module defined by Netboost.^72,82^ Functional enrichment for each protein set was assessed using Gene Ontology (GO),^73^ Kyoto Encyclopedia of Genes and Genomes (KEGG),^74^ Reactome,^75^ and WikiPathways databases. Left facing bars display pathway enrichment for proteins negatively associated with module expression. Right facing bars display pathway enrichment for proteins positively associated with module expression. The 6 proteins in each network with the highest correlation with overall network expression are designated as Hub Proteins. *Note:* * Gene shows significant differential expression in at least one brain region based on AMP-AD consortium work. **^t^** Gene shows significant differential protein expression in at least one brain region based on the AMP-AD consortium dataset (AMP-AD Sage Bionetworks Agora platform); ^★^ Gene has been nominated as an AD therapeutic target by the AMP-AD consortium.

### Mendelian randomization suggests select causal links between WMH volume and plasma proteins

To assess potential causal relationships between candidate proteins, protein modules, and WMH volume, we utilized bidirectional two-sample^31,32^ Mendelian randomization (MR).^33^ First, protein quantitative trait loci (pQTLs) for WMH-associated proteins and protein modules were identified by conducting a GWAS of plasma protein level in older adults included in the ARIC (visit 5). Plasma pQTLs identified in the INTERVAL study were used as well. pQTL instruments were identified for 10 of the 13 WMH-associated proteins and 5 of the 6 WMH-associated protein modules (*P* < 5.0 × 10^−8^). GWAS results and pQTL functional annotations, including CADD scores, are provided in **Supplementary Table 15**. Using an inverse variance weighting (IVW) method, we found no evidence for a causal relationship between proteins and WMH volume in the forward direction. However, IVW analyses found evidence for a causal link between protein module 3 (ME3) and WMH volume, though this association did not survive correction for multiple comparisons (**Table 1**; sensitivity analyses in **Supplementary Table 16**). Functional enrichment analysis for ME3 suggests involvement in complement signaling and innate immune activation (**Figure 5G**).

**Table 1.**
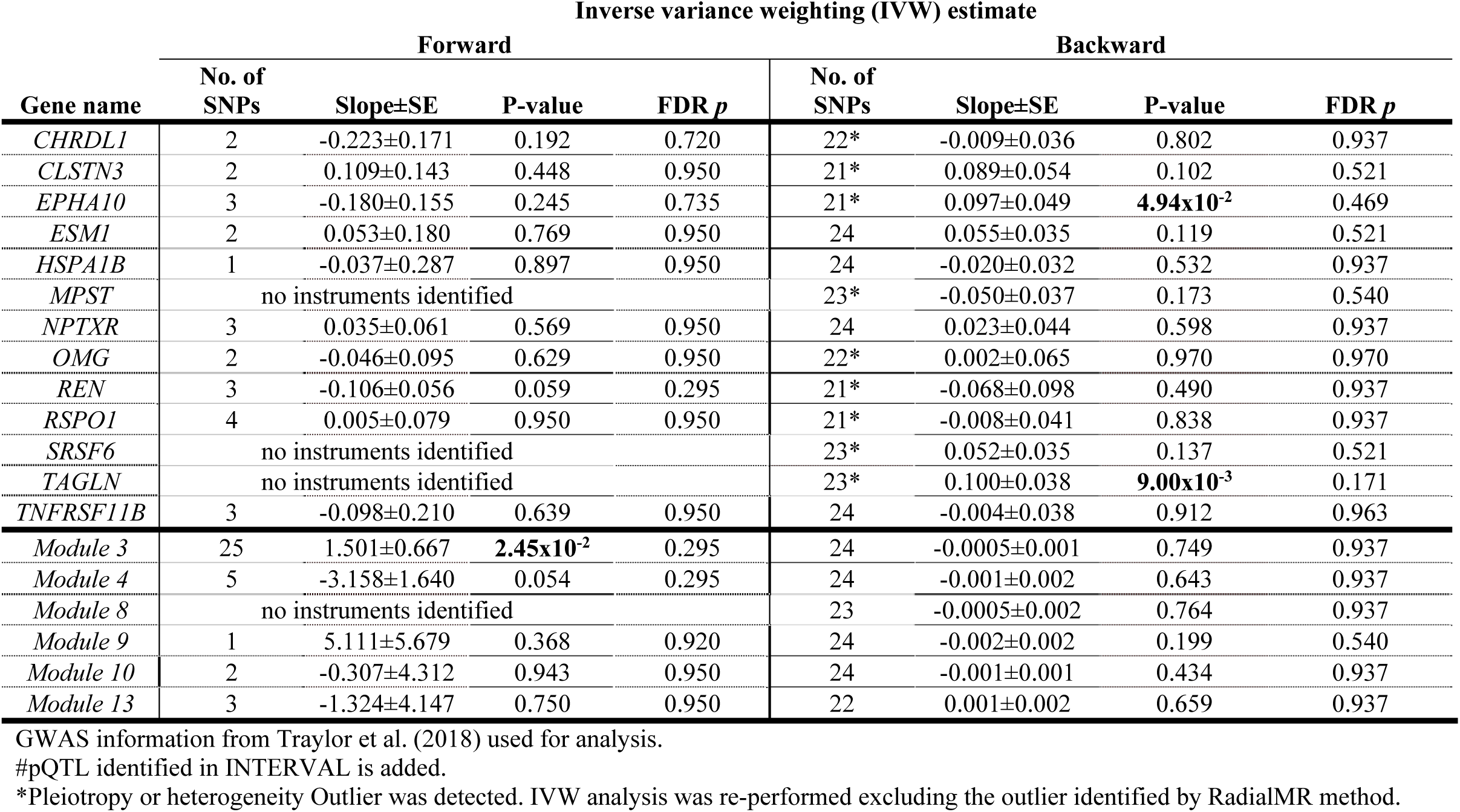
Examination of causal effects for WMH-associated proteins and protein modules using bidirectional Mendelian randomization.

Examination of the causal relationship between WMH volume and protein level (reverse direction) suggests that WMHs may be causally linked to two proteins: EPHA10 and TAGLN. Though these associations did not survive correction for multiple comparisons, suggestive evidence of this causal link between WMH volume and EPHA10 and TAGLN supports their value as potential WMH biomarkers.

### Brain tissue eQTL analysis highlights biological pathways relevant to SVD etiology

Next, we examined mRNA expression of genes in brain tissues that were associated with the identified pQTLs (a total of 1,471 genome-wide significant pQTLs). We identified 66 genes (referred to as eGenes) whose expression in brain tissues is associated with WMH-associated plasma pQTLs (**Supplementary Table 17**). Together, these genes were significantly enriched in ontologies related to lipoprotein assembly and clearance. They were also significantly enriched in GWAS of multiple traits including cerebrospinal amyloid-β_42_ and total tau levels, memory function, and myocardial infarction, mostly driven by the APOC1/C4/C2 gene region linked to WMH-associated module 13 (ME13) (**Supplementary Table 18**).

To determine whether the WMH-associated proteins and their genetic regulators (pQTLs) may affect CNS biology, we utilized a variant pathway mapping approach to identify neurobiological pathways likely altered by the genetic variation of plasma WMH-associated proteins.^35^ Incorporating GO terms for eGenes, variant pathway mapping highlighted several neurobiological pathways associated with genetically-determined plasma variation of WMH-associated proteins (**Extended Data Fig. 6; Supplementary Table 19; Supplementary Methods**). Notably, brain eGenes associated with genetic variation in OMG and NPTXR were linked to neuron projection regeneration and regulation of postsynaptic neurotransmitter receptor activity, respectively. Brain eGenes linked to WMH-associated proteins CHRDL1, RSPO1, and EPHA10 were enriched for several neural pathways, including epinephrine uptake and steroid biosynthesis. These findings further support the role of these plasma proteins as markers and/or mediators of several neurobiological processes relevant to small vessel disease.

### WMH-associated proteins predict eight-year dementia risk

Though we have identified multiple candidate biomarkers for SVD, it is also important to understand the degree to which each relates to clinically meaningful outcomes. Within a set of 4228 non-demented older adults with available SomaScan biomarkers in the ARIC study (visit 5), we examined the association of WMH-associated proteins with eight-year dementia risk (641 incident dementia cases). As illustrated in **Extended Data Fig. 7**, the majority (11 out of 13) of the WMH-associated proteins were significantly associated with dementia risk at *P*<0.05. Eight of these 11 proteins remained associated with dementia risk after FDR correction for multiple comparisons. These findings highlight the relevance of this set of SVD biomarkers to progression of clinical symptoms and dementia risk. A summary of the supportive findings for each WMH-associated protein is provided in **Extended Data Fig. 3**.

## Discussion

Given the high burden of unrecognized and untreated cerebrovascular disease and VCID among older adults, there is a critical need to better understand SVD biology and identify inexpensive and scalable biomarkers for SVD components. To this end, the current study conducted a large-scale proteome-wide analysis to investigate the plasma protein signature of WMHs, cerebral microhemorrhages, and lacunar infarcts. From a panel of nearly 5000 proteins, we identified 13 proteins and 2 protein networks that were associated with WMH volume independent of demographic and cardiovascular risk factors in a large, community-based cohort. Nine of these proteins replicated in one or more external cohort and two proteins, OMG and NPTXR, remained associated with late-life WMH volume when measured nearly two decades earlier during midlife. We demonstrated further that many of the plasma WMH-associated proteins are expressed in brain tissue wherein they are associated with pathologically defined AD, cognitive decline, and cerebral atherosclerosis. Finally, we demonstrated that 11 out of the 13 candidate WMH biomarkers are associated with risk for future dementia, underscoring their clinical relevance.

The molecular functions of the top WMH-associated proteins are together reflective of neurovascular biology. Specifically, we identified markers of endothelial dysfunction and vascular permeability (ESM1),^36^ angiogenesis (TAGLN),^37^ vascular homeostasis (MPST),^38,39^ neuronal signaling (OMG),^40,41^ astrocyte-mediated synaptogenesis (CHRDL1)^42^, and synaptic functions (NPTXR, CLSTN3, EPHA10).^43–45^ Dysregulation of these proteins in older adults may represent a disturbance of the interface between CNS cells in the brain parenchyma and its associated vasculature – termed the neurovascular unit – which has been posited as a mechanistic contributor to VCID, AD, and other neurodegenerative conditions.^46^ The neurovascular unit typifies the intimate coupling of neuronal and vascular activity, and comprises cell regulators of the blood brain barrier, including astrocytes, vascular smooth muscle cells, and endothelial cells, which can together influence neuronal integrity and function.^46,47^ Notably, 6 of the 13 candidate WMH proteins were found at measurable levels in brain tissue, and two, OMG and NPTXR, appear to be almost exclusively expressed in brain, making them attractive CNS biomarkers.

OMG, a protein expressed primarily by oligodendrocytes and astrocytes, functions in the formation and maintenance of myelin sheaths and negatively regulates neurite outgrowth.^40,41^ Given that OMG has been identified as a myelin-associated inhibitor of axonal growth and implicated in conditions known to affect myelin integrity (e.g., multiple sclerosis), this protein may serve as a general indicator of white matter health.^40,48,49^ We suspect low plasma levels of this brain-derived protein may reflect reduced brain OMG expression as a result of white matter lesions. Alternatively, individuals with low OMG expression may be more vulnerable to white matter lesions. Another possibility is that lower plasma OMG may paradoxically reflect elevated OMG expression in brain tissue. In support of this idea, higher level of OMG in brain tissue was associated with greater levels of cerebral atherosclerosis and cognitive decline,^27,28^ whereas low level of this protein in plasma was associated with greater WMH volume in multiple cohorts and greater risk for subsequent dementia. NPTXR, on the other hand, is a regulator of neurotransmitter receptor activity at the synapse and a receptor for AD biomarker, NPTX2.^50^ Knockdown studies of this type II transmembrane protein have demonstrated decreased assembly and function of excitatory and inhibitory postsynaptic processes.^43^ NPTXR modulates synaptic function^43,51^ by coordinating glutamate receptors and regulating the behavior of secreted pentraxins, including neuronal pentraxin 1, which mediates Aβ-induced synapse loss and neuronal apoptosis.^52^ Several studies have identified NPTXR as a protein which declines in a stepwise manner as disease progresses from mild cognitive impairment (MCI) to moderate AD, underscoring the utility of this protein for AD staging.^53^ We extend these findings by demonstrating an inverse association between *plasma* NPTXR levels and WMH volume and subsequent dementia risk. NPTXR’s function as a WMH biomarker is further supported by the inverse association between brain levels of this protein and cerebral atherosclerosis and cognitive decline.^27,28^

As part of the current study, we used genetic information to test the causal link between candidate proteins and WMH volume. Although our findings do not causally implicate any of the 13 candidate proteins in WMH development, we did find evidence for a causal relationship between a network of complement/immune signaling proteins (ME3) and WMH volume. This protein network includes as a hub protein VTN (a.k.a. complement S-protein), which is a cell adhesion and migration glycoprotein that has been nominated by AMP-AD as a therapeutic target for AD. Our results suggest complement pathways (C3 and C5) or drivers of this immune/complement protein network, such as VTN, play a mechanistic role in WMH development and could function as therapeutic targets for VCID. We also note that the expression of a module characterized by interleukin-11 signaling, spliceosome function, and SARS-CoV-2 pathways (NRP1) was broadly associated with all measured forms of cerebral SVD. Altered expression of this network may be an indicator of white matter alterations in the context of dementia, given that (1) three of the hub proteins – EPHA10, CLSTN3, and HSPA1B – were among the top proteins associated with 25-year dementia risk in a recent ARIC study, (2) nearly all ME10 hub proteins were differentially expressed in AD brains at the level of protein or transcript, and (3) interleukin-11 has been found to regulate inflammatory demyelination.^54^

The genetic architecture of cerebral SVD overlaps considerably with cardiometabolic disorders, and this overlap is to an extent also reflected in the plasma proteome. One GWAS found that over half of WMH risk variants were also associated with blood pressure or known vascular risk factors such diabetes and LDL cholesterol.^55^ While only two WMH-associated proteins identified in the current study have been genetically linked to blood pressure (ESM1, REN), we did find that half the genes coding for WMH-associated proteins have been implicated in at least one form of cardiometabolic dysfunction. Pathway analyses show distinct biology linked to distinct forms of SVD, but also suggest the involvement of coagulation, atherosclerosis, and cholesterol regulation (LXR/RXR) – which may be important in myelin maintenance and repair^56^ – in multiple forms of SVD. These results also suggest an association between lacunar infarcts and altered immune pathways (NF-kB signaling, acute phase response, IL-15, and IL-6 signaling), a finding that was corroborated orthogonally by our network analyses and supported by extant literature linking inflammation to lacunar infarcts.^57,58^ Future studies will need to parse out whether peripheral immune aberrations are a cause or merely a consequence of lacunar infarcts.

This study, one of the earliest large-scale plasma proteomic analyses of cerebral SVD, has multiple strengths, including use of several large multi-racial community samples, measurement of plasma proteins at multiple time-points, robust measurement and adjustment for potential confounders, and the use of network and causal methodology. However, these results should be interpreted in the context of several limitations. First, the ARIC study sample – the cohort within which the discovery analyses were conducted – is comprised of Black and White participants from four communities within the U.S. Though the external replication analyses support the generalizability of these results, our findings may not be generalizable to non-Black and non-White groups within the U.S. or those living outside the U.S. Second, several factors including MRI strength, signal-to-noise ratio (SNR), as well as detection and quantification methods can impact the sensitivity of lesion detection and quantification, thus impacting the prevalence of neuroimaging characteristics in the cohorts included in this study. Third, the power to detect protein associations was likely limited in our analysis of lacunar infarcts and CMHs given that these traits were examined as binary outcomes. Lastly, though the SomaScan Proteomic Assay measures an extensive sample of the human proteome, it does not quantify levels of all proteins found in plasma. Despite these limitations, this large-scale proteomic analysis within a longitudinal community-based sample identified several proteins and pathways that may be particularly important in the development of SVD.

## Methods

### Study Design and Participants

This study utilized data from the Atherosclerosis Risk in Communities study, a community-based cohort study that enrolled 15,792 participants from four U.S. communities: Washington County, Maryland; Forsyth County, North Carolina; northwestern suburbs of Minneapolis, Minnesota; and Jackson, Mississippi between 1987 and 1989.^59^ Study visits of direct relevance to the present analysis include visit 3 (1993-1995) and visit 5 (2011-2013). Throughout the study there was continuous surveillance for hospitalizations and death (for study timeline, see **Extended Data Figure 1**). Study protocols were approved by Institutional Review Boards at all participating centers: University of North Carolina at Chapel Hill, Chapel Hill, NC; Wake Forest University, Winston-Salem, NC; Johns Hopkins University, Baltimore, MD; University of Minnesota, Minneapolis, MN; and University of Mississippi Medical Center, Jackson, MS. All participants gave written informed consent at each study visit, and proxies provided consent for participants who were judged to lack capacity.

As displayed in **Figure 1**, the primary analysis utilized SomaScan Multiplexed Proteomic technology to examine the association of plasma protein levels at ARIC visit 5 (late-life) with white matter hyperintensity volume, the presence of lacunar infarcts, and the presence of cerebral microhemorrhages also measured at the same visit. Exclusion criteria included non-Black/non-White race, non-white participants in Washington County and Minnesota, or missing proteomic, neuroimaging, or covariate data (**Extended Data Figure 1**). After participant exclusion, 1508 participants were included in the primary analysis. As part of an internal replication analysis, we measured candidate proteins at an earlier wave of the ARIC study (visit 3; 1993-1995) using the same SomaScan platform. Using the methods described below, these candidate proteins were then related to late-life SVD variables (see **Extended Data Figure 1**). A total of 1462 participants were included in this internal replication analysis.

### Protein Measurement

Plasma was collected at each ARIC study site at visit 3 and visit 5. Consistent with standardized protocol, plasma was frozen at 80°C and shipped on dry ice to the ARIC central laboratory where it remained continuously frozen until it was aliquoted into barcoded microtiter plates. Plates were then sent to SomaLogic for quantification (SomaLogic, Inc, Boulder, Colorado). SomaLogic’s SomaScan platform was used to measure the relative concentration of 5,284 modified aptamers (SOMAmer reagents or ‘SOMAmers’) using a Slow Off-rate Modified Aptamer (SOMAmer)-based capture array.^60^ The SOMAmer method utilizes modified aptamers—short single strands of DNA with chemically modified nucleotides—as protein binding reagents which can be uniquely identified and quantified using DNA detection technology. This SomaScan v.4 platform used in this study, which has been detailed previously,^24,61,62^ provides relative concentration of approximately 5,000 plasma proteins or protein complexes. The median intra-and inter-run coefficients of variation for the SomaScan platform have been reported as approximately 5%. The median intraclass correlation coefficient has been reported as approximately 0.90.^63,64^ SomaScan assay performance characteristics have been detailed previously.^65–68^ After performing quality control steps outlined in the **Supplementary Materials**, a total of 4,877 SOMAmers measuring 4,697 unique proteins or protein complexes were analyzed. Proteins were log2 transformed and outliers >5 SD away from the mean were winsorized. Inter-assay CV_BA_s and reliability coefficients for SOMAmers associated with WMH volume are provided in **Supplementary Table 20**.

### Covariate assessment

The following participant variables were reported at ARIC visit 1: sex (male/female), race (Black/White), education (less than high school/high school, general education diploma [GED], or vocational school/at least some college). Due to the confounding nature of race and study center variables in this study, a combined race-center variable was created as follows: White-Washington County, white-Forsyth County, Black-Forsyth County, white-Minneapolis, or Black-Jackson. APOE was assessed using the TaqMan assay (Applied Biosystems, Foster City, CA) and was coded as 0 APOEε4 alleles, ≥1 APOEε4 alleles, or missing. Remaining covariates were captured at visit 5 or visit 3, i.e., the visit concurrent with proteomic measurement. Estimated glomerular filtration rate (eGFR) was calculated using serum creatinine and demographic characteristics.^69^ Missing eGFR at visit 3 was imputed from visit 2 calculations. Body mass index (BMI) was calculated using height and weight (kg/m2). At visit 5, diabetes was defined based on HbA1c level of 6.5% or greater, current use of diabetes medication, or self-report of diagnosis by a physician. At visit 3, diabetes was defined as a fasting glucose ≥126 mg/dL or non-fasting glucose ≥200 mg/dL, current use of diabetes medication, or a self-report of diagnosis by a physician. Hypertension was defined utilizing the mean of the last two of three blood pressure measurements as a systolic blood pressure >140 mm Hg, a diastolic blood pressure >90 mm Hg, or use of hypertensive medication. Smoking was assessed as a binary variable (yes/no) based on self-report of current smoking status.

#### Brain Magnetic Resonance Imaging (MRI)

As part of the ARIC Neurocognitive Study (NCS), 3T MRIs were conducted in approximately 2,000 participants at visit 5. The MRI selection criteria are described in the **Supplemental Materials**. The acquisition sequence for the ARIC visit 5 MRI has been described previously^14^. At each ARIC site, a common set of sequences were performed for all participants: MP-RAGE, Axial T2*GRE, Axial T2 FLAIR, and Axial DTI.

##### White Matter Hyperintensity (WMH) Volume

WMH volume (mm^3^) was assessed quantitatively from FLAIR images using a computer-aided segmentation program to assess the total volumetric burden.^70^ All analyses of WMH volume were adjusted for total intracranial volume. WMH volume was log-transformed to adjust for skewness. *Cerebral microhemorrhages (CMHs) and lacunar infarction.* CMHs and lacunar infarcts were identified by trained imaging technicians and confirmed by radiologists; this has been previously described.^71^ Lacunar infarcts were defined as a hyperintense subcortical lesion with a dark center (≥3mm and ≤15mm in size) within the white matter, infratentorial, or central grey/capsular regions that is distinguishable from perivascular space. CMHs were defined using a T2* GRE MRI sequence and characterized by location: subcortical CMHs and lobar CMHs.

##### Identification of plasma proteins associated with SVD neuroimaging variables

Separate multivariable linear regression models were used to examine the association between the level of each protein and WMH volume. Separate multivariable logistic regression models were used to assess the relationship between protein levels and subcortical CMHs, lobar CMHs, and lacunar infarcts. Analyses were first adjusted for age at sample acquisition, sex, education, race-center, intracranial volume, and kidney function, defined as eGFR-creatinine (model 1). Second, analyses were additionally adjusted for cardiovascular risk factors, i.e. BMI, hypertension, diabetes, and smoking status (model 2). The second model was used for all primary analyses. An FDR corrected *P<*0.05 threshold was used to identify candidate proteins in proteome-wide analyses. Sensitivity analyses examined the effect of including (1) *APOE*ε4 status and (2) prevalent stroke covariates, in addition to (3) using sampling weight to account for selection into the ARIC MRI study. To identify the biological pathways underlying the differential expression of SVD-associated proteins, we conducted a canonical (biological) pathway analysis using the Ingenuity Pathway Analysis (IPA) bioinformatic platform (see **Supplementary Methods**).

Candidate proteins significantly associated with SVD neuroimaging variables in the primary analysis were measured using the same SomaScan platform 18 years earlier at ARIC visit 3. These proteins were related to the same late-life SVD neuroimaging variables using an approach similar to that described above. Specifically, we used linear and logistic regression models adjusted for visit 3 age, other demographic variables (sex, race-center, education), kidney function defined as eGFR-creatinine, and cardiovascular risk factors measured at visit 3 (BMI, diabetes, hypertension, and smoking status).

##### Identification of SVD-associated protein networks

Using the 4877 SomaScan proteins measured at ARIC visit 5, we used the Bioconductor R package Netboost v2.1.1^29,72^ to identify networks of co-expressed proteins based on the sparsity of protein networks. Using the steps described in the **Supplementary Methods**, we identified 15 protein modules. The first principal component score for the rank matrix of each protein module, referred to as the module eigenprotein (ME), was calculated to quantify person-specific protein module expression. The protein-module assignments and the correlations between each protein and the calculated ME (module membership) are provided in **Supplementary Table 14**. Module membership values were used to define hub proteins for each module. The ME values were examined as exposure variables and related to MRI-defined SVD in separate multivariable linear and logistic regression models using covariates defined in fully-adjusted model 2 described above. SVD-associated protein networks were characterized for biological significance using the g:Profiler toolkit gene set enrichment analysis which applied Gene Ontology (GO),^73^ Kyoto Encyclopedia of Genes and Genomes (KEGG),^74^ Reactome,^75^ and WikiPathways^76^ databases.

##### Mendelian Randomization Analysis

A bi-directional two-sample Mendelian randomization (MR) design^31^ was used to determine whether there is evidence of a causal link between WMH-associated proteins and WMH volume. The methods used to detect instrumental variables (IVs) are defined in the **Supplementary Methods**. Multi-IV MR analyses were performed when proteins had at least three IVs. Otherwise, we performed single IV MR using an inverse-variance weighted (IVW) method. The IVW method was used as the primary approach for multi-IV MR.^17^ These analyses were supplemented by several sensitivity analyses using complimentary approaches, including MR-Egger^18^, weighted-median method^19^ and CONtamination MIXture^20^, which are robust to violation of the assumptions. MR assumptions were also evaluated. The strength of a set of IVs was tested using the Staiger-Stock rule.^21^ Horizontal pleiotropy was evaluated using the Egger intercept test^23^ and the Mendelian Randomization Pleiotropy RESidual Sum and Outlier test^24^. Heterogeneous effects among causal estimates were examined using Cochran’s heterogeneity Q for IVW and Egger methods^25^. Outlier instruments were detected using radial plots constructed using the RadialMR^26^ (v.0.4) R package. If an outlier was detected, MR analyses were performed again after excluding outliers. The two-sided *P*-values derived from primary IVW analyses were adjusted for multiple comparisons using FDR. All analyses were performed using TwoSampleMR^27^ (v.0.5.5) and MendelianRandomization^28^ (v.0.5.0) R packages.

##### External replication of SVD-associated proteins

To assess the reproducibility and generalizability of the protein associations with SVD metrics in the ARIC cohort, we conducted protein-MRI analyses using three external cohort studies: Baltimore Longitudinal Study of Aging (BLSA), Generation Scotland (GS), and the Cardiovascular Health Study (CHS). We examined candidate proteins in relation to WMH volume (BLSA, GS, CHS), white matter grade (CHS), and total white matter volume (BLSA). A description of each cohort and the accompanying analyses are provided is provided in the **Supplementary Methods**.

##### SVD-associated proteins and dementia risk

To determine the clinical utility of SVD-associated proteins, we related candidate proteins to dementia risk within the ARIC cohort. Using methods consistent with those employed previously in a large-scale proteomic analysis of dementia risk within the ARIC study, we related candidate proteins measured at ARIC visit 5 in an overlapping cohort of non-demented participants to dementia risk occurring through ARIC visit 7 (2018-2019). The full exclusion and inclusion criteria for this analysis is provided in **Extended Data Figure 1**. ARIC participant characteristics stratified according to late-life baseline (2011-2013) cognitive status are included in **Supplementary Table 22**. We used Cox proportional hazard models adjusted for demographic variables (age, sex, race-center, education), *APOE*ε4 status, kidney function defined as eGFR-creatinine, and cardiovascular risk factors (BMI, diabetes, hypertension, and smoking status). The methods used for dementia classification in ARIC have been defined in detail elsewhere.^77^

## Data availability

All data generated in this study are either included in this article (and its Supplementary Information), available on reasonable request, or are available in an online public database. Pre-existing data access policies for each of the parent cohort studies (ARIC, CHS, BLSA, GS) specify that research data requests can be submitted to each steering committee; these will be reviewed for confidentiality or intellectual property restrictions and will not unreasonably be refused. Individual level patient or protein data may further be restricted by consent, confidentiality or privacy laws/considerations. These policies apply to both clinical and proteomic data. Tissue-specific gene expression data is available at https://www.gtexportal.org/home/. Tissue-specific protein expression data is available at https://www.proteinatlas.org/. Brain cell-specific gene expression data is available at http://www.brainrnaseq.org/. Brain gene expression date were derived from the Brain eQTL Almanac (BRAINEAC; http://www.braineac.org/) and the Functional Mapping and Annotation of Genome-Wide Association Studies105 (FUMA) platform (https://fuma.ctglab.nl/). eQTL gene enrichment was performed using data from the Molecular signatures (MsigDB; http://www.broadinstitute.org/msigdb) and the NHGRI-EBI Catalog of published genome-wide association studies (GWAS Catalog; https://www.ebi.ac.uk/gwas/). Functional enrichment of protein networks was conducted using the g:Profiler web tool https://biit.cs.ut.ee/gprofiler/gost.

## Code availability

All software used in this study are publicly available: R version 4.1.2 (https://www.r-project.org/); Ingenuity Pathway Analysis (https://digitalinsights.qiagen.com/products-overview/discovery-insights-portfolio/analysis-and-visualization/qiagen-ipa/); EplainBio (http://www.explainbio.com/); GraphPad Prism 9.4.1. (https://www.graphpad.com/scientific-software/prism/). The code used in this study can be made available from the corresponding author on reasonable request.

## Funding & Acknowledgements

The Atherosclerosis Risk in Communities Study is carried out as a collaborative study supported by National Heart, Lung, and Blood Institute contracts (HHSN268201700001I, HHSN268201700002I, HHSN268201700003I, HHSN268201700004I, HHSN268201700005I). Neurocognitive data is collected by U01 2U01HL096812, 2U01HL096814, 2U01HL096899, 2U01HL096902, 2U01HL096917 from the NIH (NHLBI, NINDS, NIA and NIDCD), and with previous brain MRI examinations funded by R01-HL70825 from the NHLBI. The authors thank the staff and participants of the ARIC study for their important contributions. Funding was also supported by R01HL087641, R01HL059367 and R01HL086694; National Human Genome Research Institute contract U01HG004402; and National Institutes of Health contract HHSN268200625226C. Infrastructure was partly supported by Grant Number UL1RR025005, a component of the National Institutes of Health and NIH Roadmap for Medical Research. SomaLogic Inc. conducted the SomaScan assays in exchange for use of ARIC data. This work was supported in part by NIH/NHLBI grant R01 HL134320. Reagents for GDF15 assays were donated by the Roche Diagnostics Corporation. The Cardiovascular Health Study was supported by contracts HHSN268201200036C, HHSN268200800007C, HHSN268201800001C, N01HC55222, N01HC85079, N01HC85080, N01HC85081, N01HC85082, N01HC85083, N01HC85086, 75N92021D00006, and grants U01HL080295 and U01HL130114 from the National Heart, Lung, and Blood Institute (NHLBI), with additional contribution from the National Institute of Neurological Disorders and Stroke (NINDS); additional support was provided by N01HC15103 and R01AG023629 from the National Institute on Aging (NIA). In addition, this work was supported, in part, by the Intramural Research Program of the National Institute on Aging-NIH. M.R.D., G.N.D., Z.P., and K.A.W. are supported by the National Institute on Aging Intramural Research Program. R.F.G. is supported by the National Institute of Neurological Disorders and Stroke Intramural Research Program. A.E.F. is supported by K01AG071689. P.S. was supported by the German Research Foundation (DFG) Project-ID 431984000 – CRC 1453 NephGen, Project-ID 192904750 – CRC 992 Medical Epigenetics, and by the DFG grant SCHL 2292/1 (Walter Benjamin Fellowship). T.M.H. is supported by the following grants from the National Institutes of Health: P30AG072947, R01AG054069, R01AG058969, RF1NS110043, U01HL096812, R01HL153191, R01AG070867, R01AG071032, R01AG072634, R01AG068629, R01HL158622, R01AG070881, R01AG067557, R21AG075291, and R01AG074971. P.L.L. is supported by NIH/NHLBI K24 HL159246. M.F. was supported in part by NIA grants U01AG052409 and U01AG058589. L.S. is funded by the Virtual Brain Cloud from European commission (grant no. H2020-SC1-DTH-2018-1).

## Author contributions

G.T.G., M.F., T.M.H., R.F.G., T.H.M, J.C. (Josef Coresh), and K.A.W. conceptualized and designed the study. W.T.L., B.M.P., A.J.N., N.J.B., C.R.J., R.F.G., T.H.M., and J.C. (Josef Coresh) contributed to the data acquisition. G.T.G., L.S., A.E.F., J.C. (Jingsha Chen), Y.Y., M.F., M.R.D., Z.P., G.N.D., P.S., W.T.L., B.M.P., and K.A.W. analyzed the data. G.T.G., L.S., A.E.F., M.F., M.R.D., A.T., P.L.L., R.F.G., T.M.H., J.C. (Josef Coresh) and K.A.W. contributed to the interpretation of data. G.T.G., L.S., A.E.F., M.F., M.R.D., Z.P., A.T., P.S., W.T.L., R.K., M.S., B.M.P., A.J.N., N.J.B., R.F.G., P.L.L., C.R.J., K.J.S., T.H.M., T.M.H., and J.C. (Josef Coresh) drafted and provided substantial revision of the manuscript.

---

**Extended Data Figure 1.**
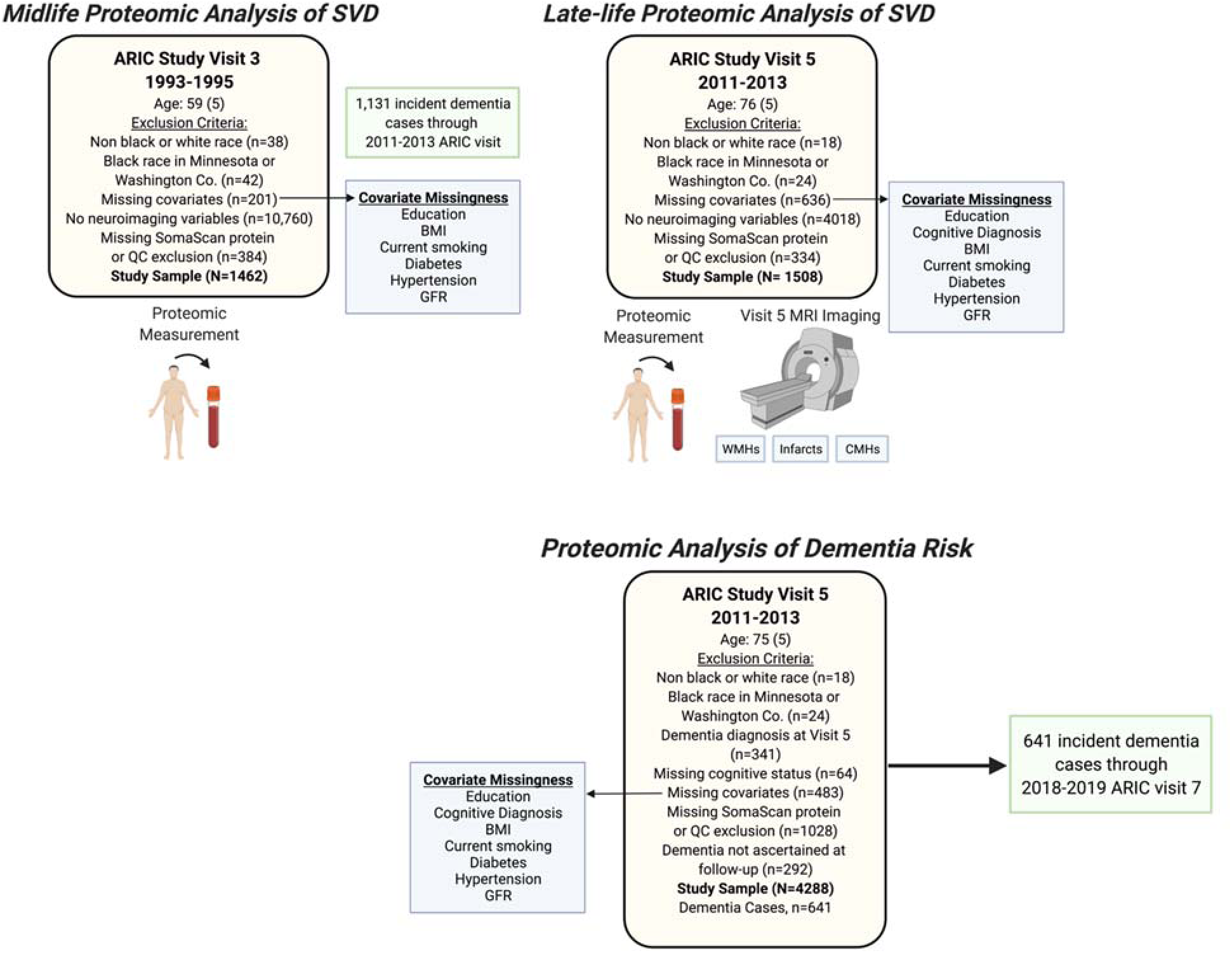
ARIC Study Timeline.

**Extended Data Figure 2.**
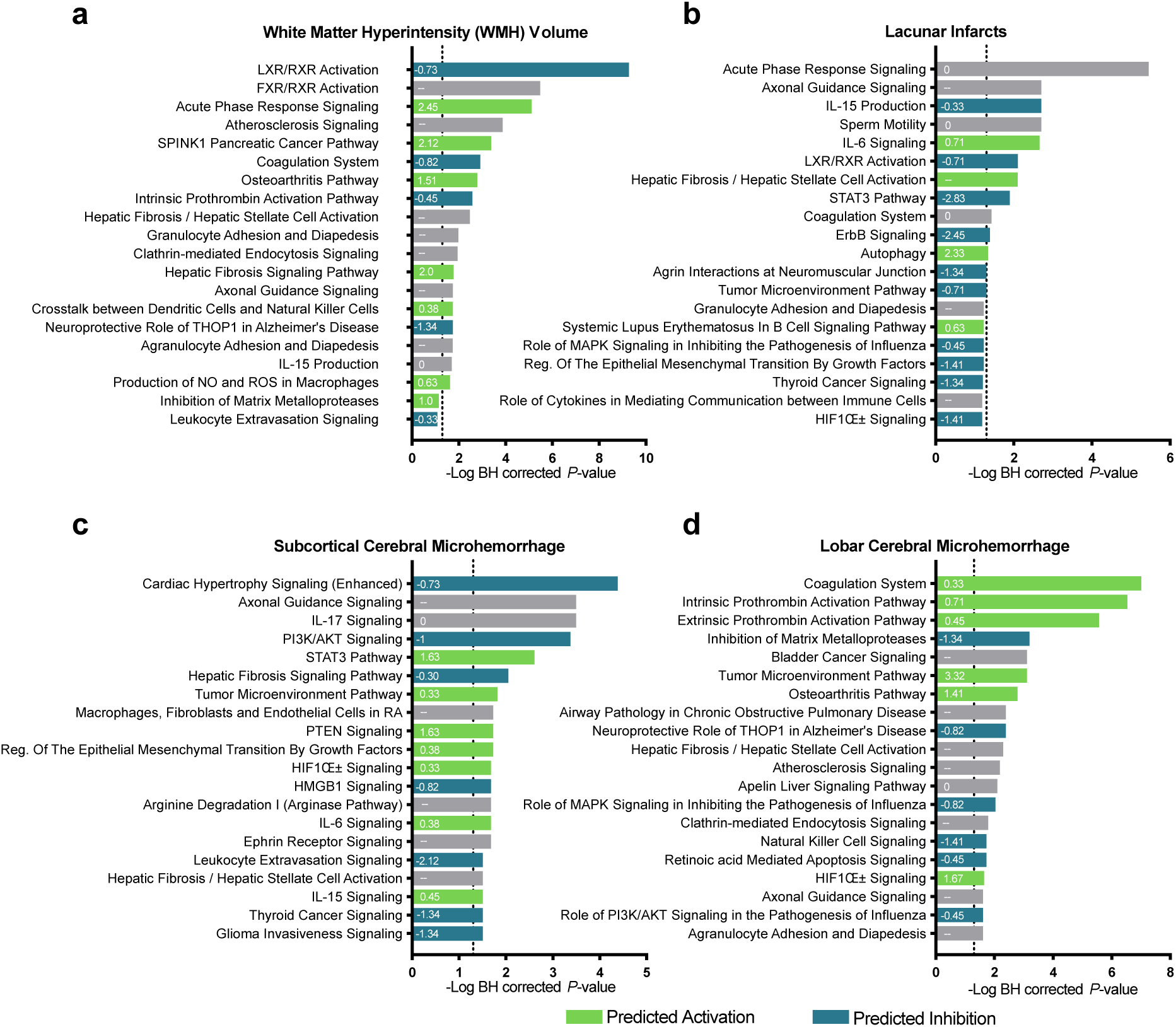
Enriched biological pathways for proteins associated with cerebral small vessel disease. For each small vessel disease characteristic—white matter hyperintensities, lacunar infarcts, subcortical and lobar cerebral microhemorrhages—the top 25 canonical pathways, identified using IPA, are displayed. The threshold for statistical significance, defined as an FDR-corrected P<0.05, is represented by the vertical dotted line. The Z-score indicating the predicted degree of activation or inhibition for each pathway is displayed within each bar. For bars with no Z-scores, the extent of activation could not be predicted. For gray bars, the direction of activation could not be predicted. These results incorporate the full gene list in the IPA database as the reference gene set.

**Extended Data Figure 3.**
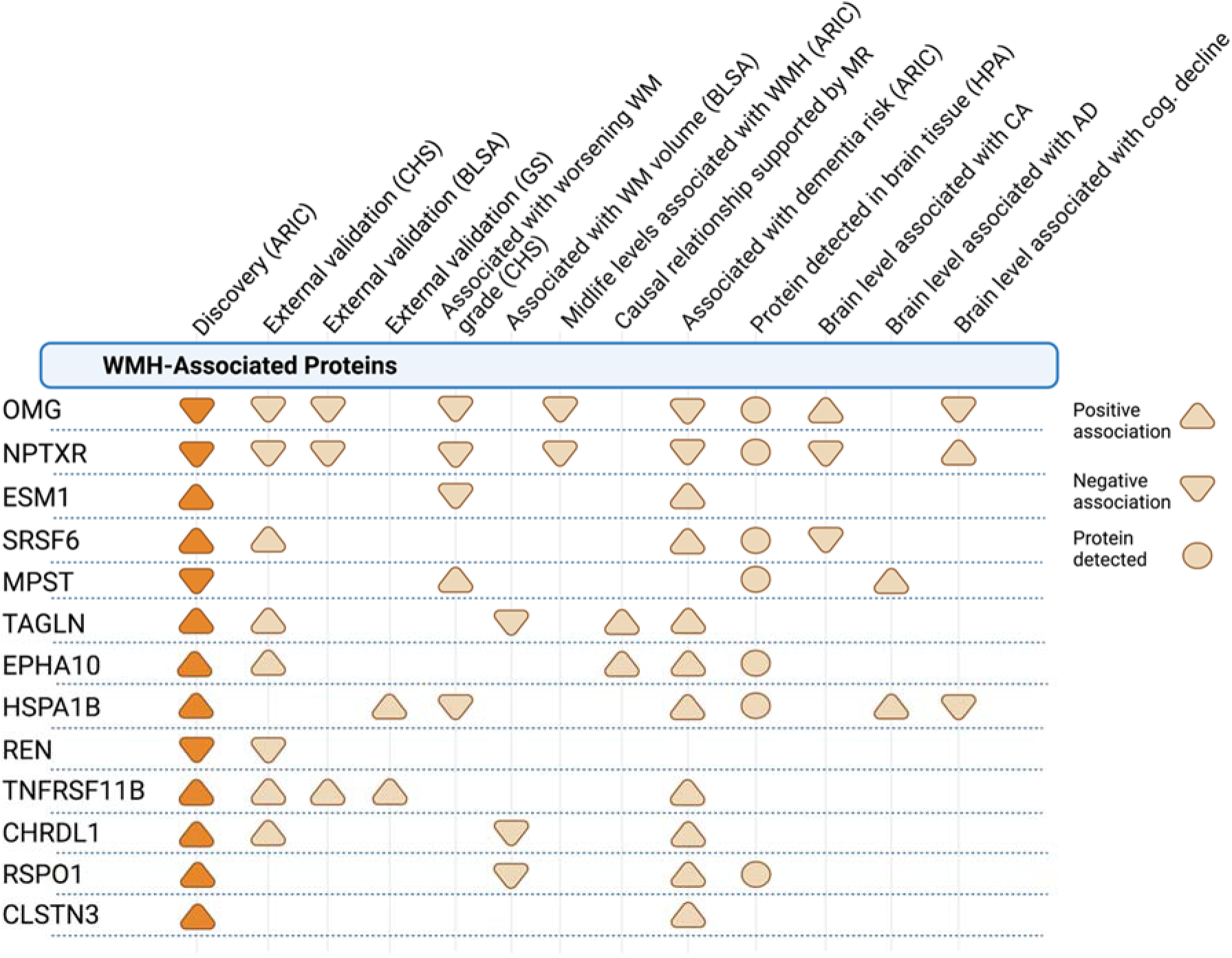
Summary of supportive evidence for WMH-associated proteins.

**Extended Data Figure 4.**
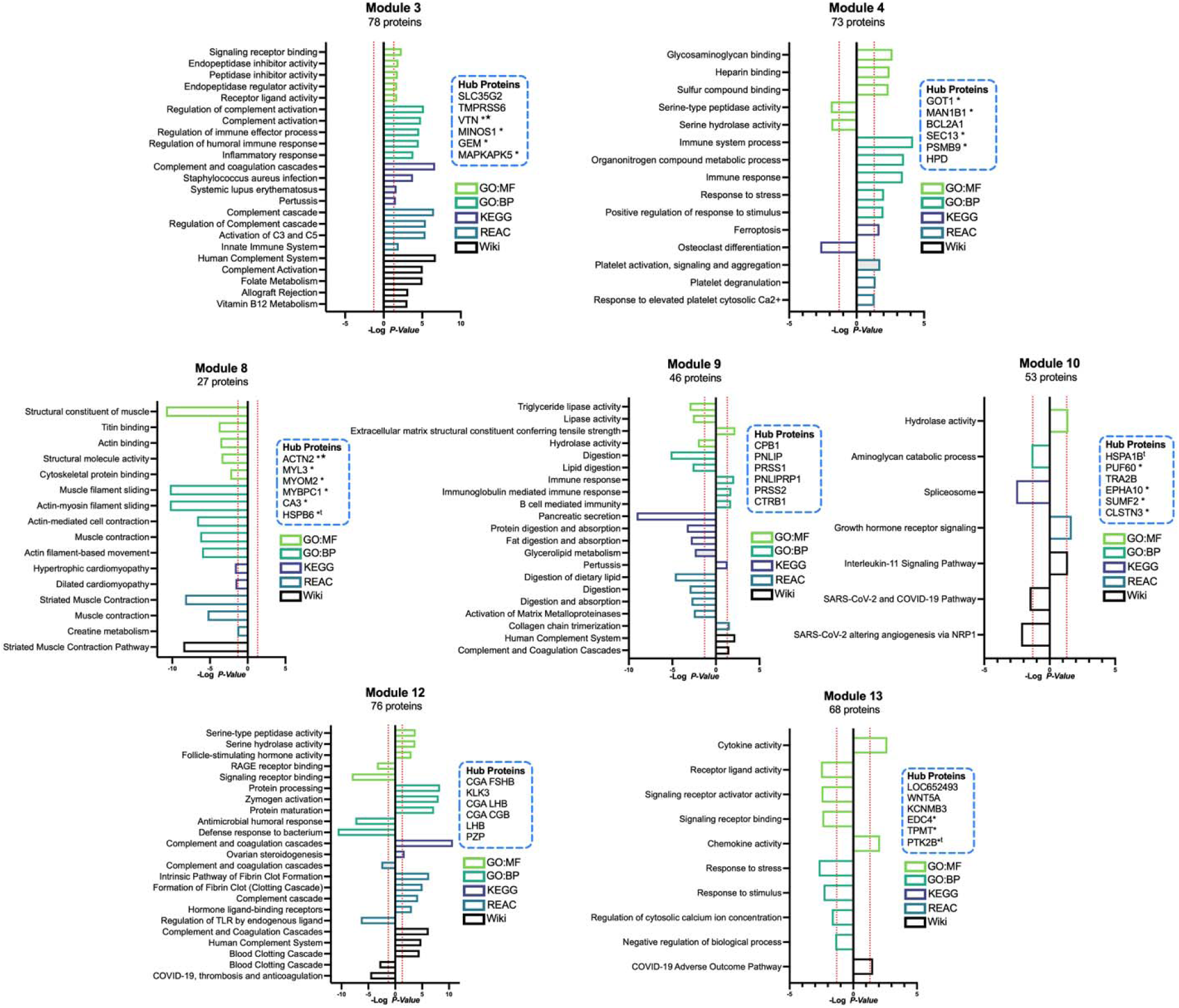
Functional profiling and tissue enrichment analyses for all modules associated with at least one cerebral small vessel disease characteristic. Gene set enrichment (using the g:Profiler toolkit) of each WMH-associated protein module was defined by Netboost.^72,82^ Functional enrichment for each protein set was assessed using Gene Ontology (GO),^73^ Kyoto Encyclopedia of Genes and Genomes (KEGG),^74^ Reactome,^75^ and WikiPathways databases. The left facing bars indicate pathway enrichment for proteins negatively associated with module expression. Right facing bars indicate pathway enrichment for proteins positively associated with module expression. The 6 proteins in each network with the highest correlation with overall network expression are designated as Hub Proteins. Modules 9, 10 and 12 were associated with at least one SVD variable at a Bonferroni corrected threshold (p <0.003). *Note:* * Gene shows significant differential expression in at least one brain region based on AMP-AD consortium dataset (AMP-AD Sage Bionetworks Agora platform). ^t^ Gene shows significant differential protein expression in at least one brain region based on AMP-AD consortium dataset (AMP-AD Sage Bionetworks Agora platform); ^★^ Gene has been nominated as an AD therapeutic target by the AMP-AD consortium.

**Extended Data Figure 5.**
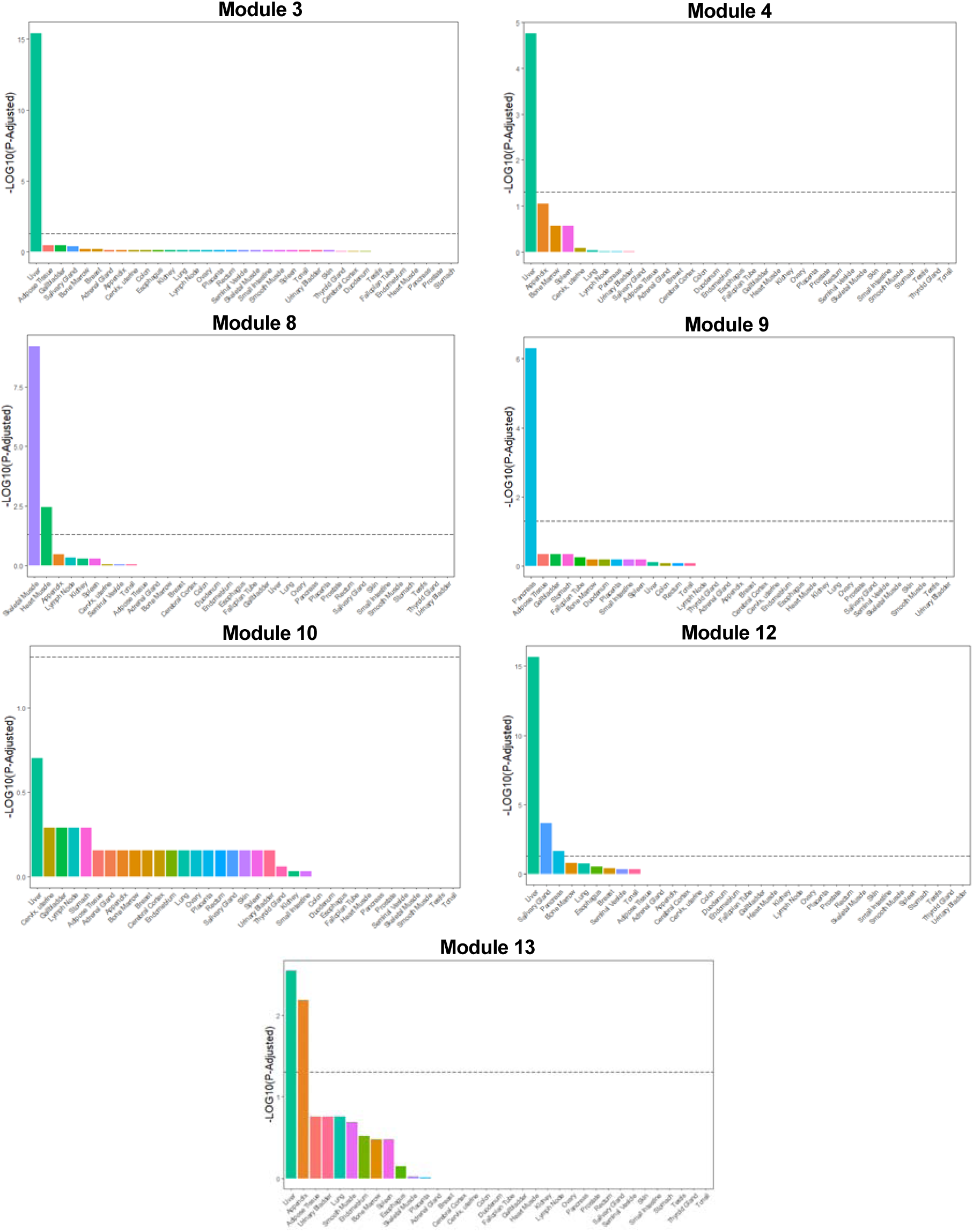
Tissue enrichment for proteins within WMH-associated modules. Tissue enrichment analysis based on tissue-specific genes from the Human Protein Atlas (HPA) TissueEnrich^83^ gene enrichment analysis package. Statistical significance (P<0.05 after FDR correction for multiple comparisons) is noted by the horizontal dashed line.

**Extended Data Figure 6.**
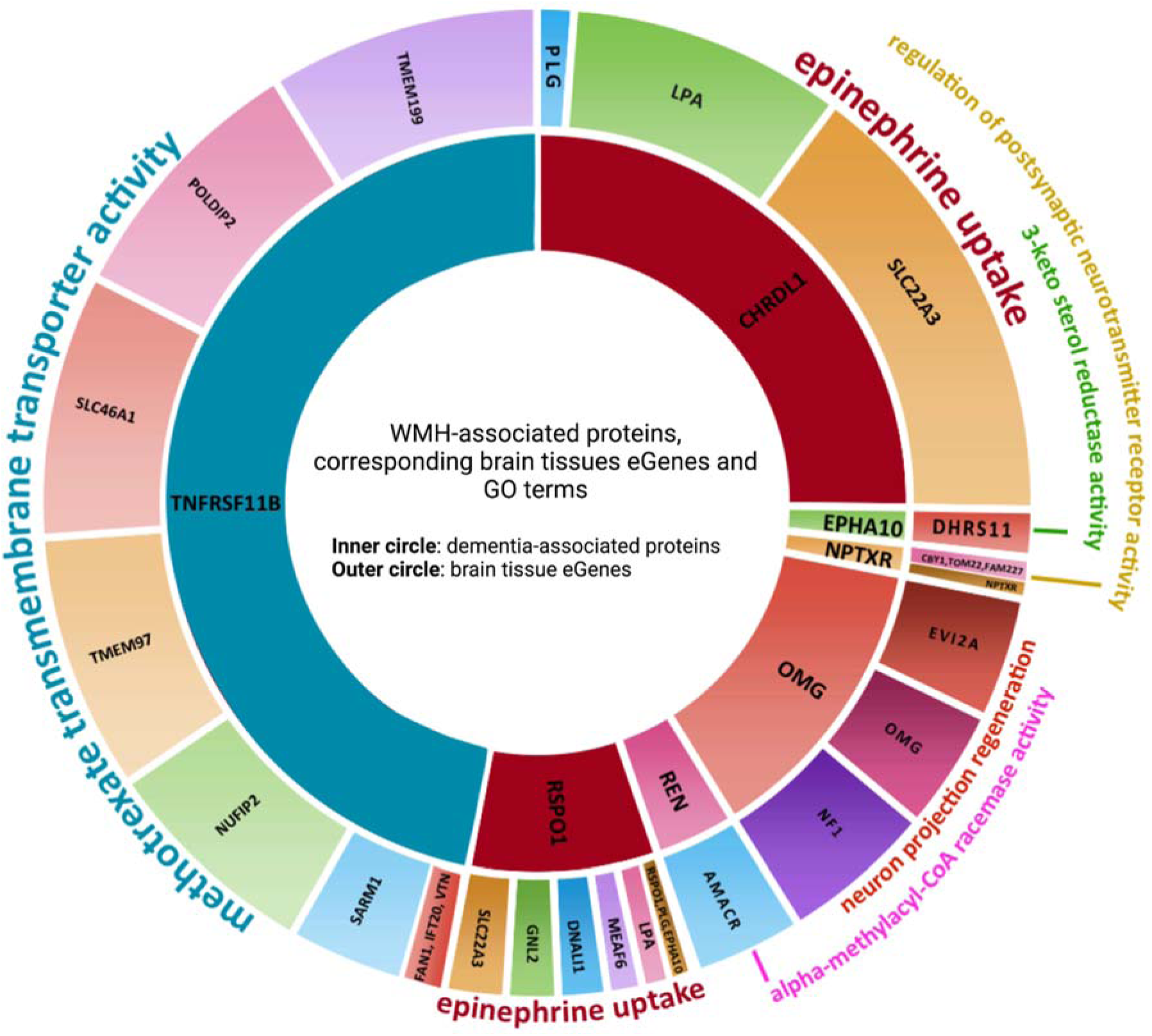
Brain tissue gene expression and biological pathways associated with genetically determined WMH-associated plasma protein level. Results of Variant-to-Pathway Mapping (VPM) analyses depicting GO Terms most likely to be associated with the set of brain tissue eGene(s) linked to each WMH-associated protein. Go terms are listed outside the circle in colored text corresponding to their respective WMH-associated protein. This analysis considered Gene Ontology (GO) terms for Biological Processes and/or Molecular Functions supported by peer-reviewed documentation. The size of the inner circular segment, listing WMH-associated proteins, is defined by the magnitude of the association between the WMH-associated protein pQTLs and the GO term linked to the set of eGenes. The size of the outer circular segment is defined by the weighted proportion of eQTLs associated with each eGene.

**Extended Data Figure 7.**
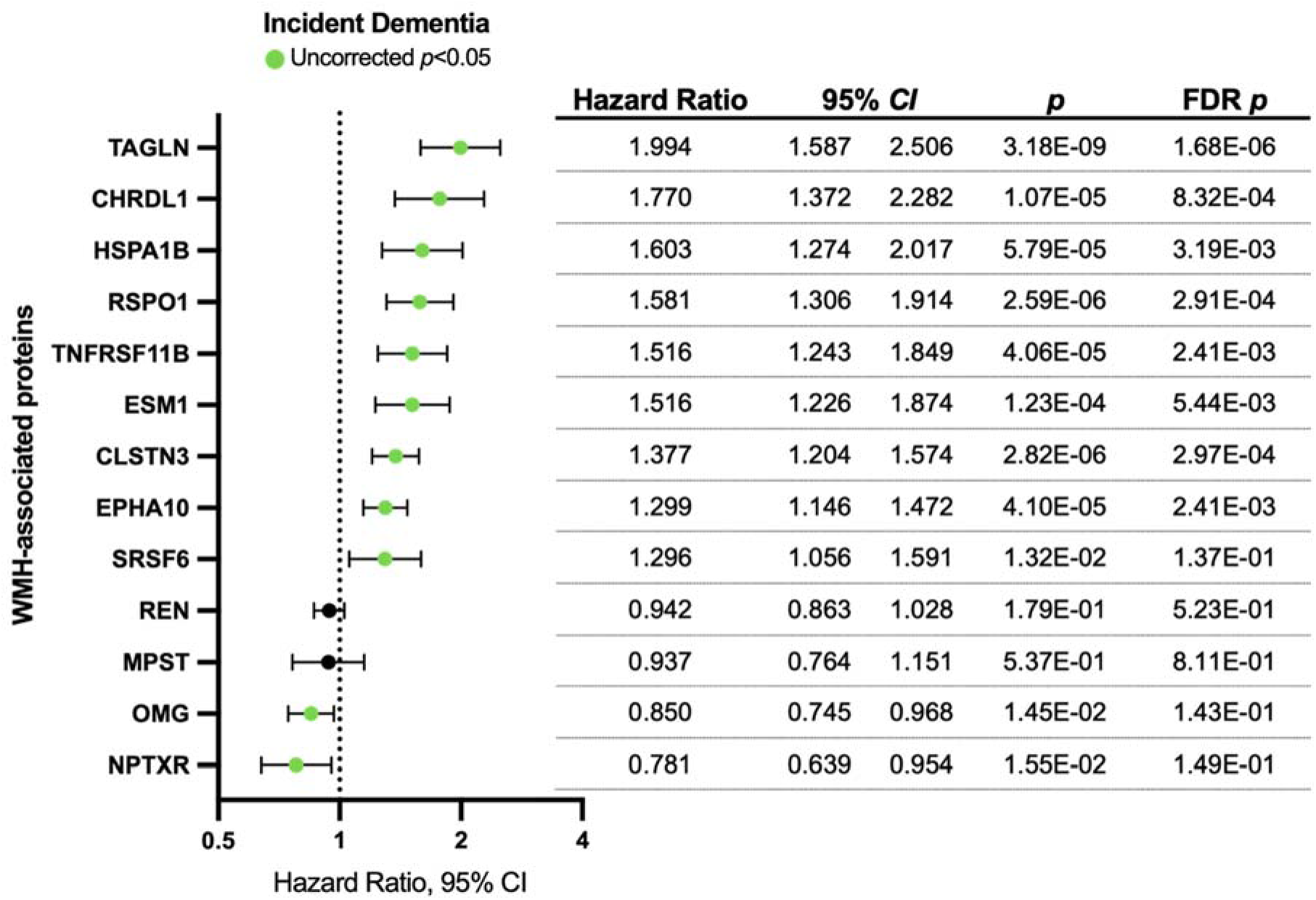
Relationship between WMH-associated proteins and incident dementia in the ARIC Study. Hazard ratios were derived from Cox proportional hazard models adjusted for demographic variables (age, sex, race-center, education), *APOE*ε4 status, kidney function defined as eGFR-creatinine, and cardiovascular risk factors (BMI, diabetes, hypertension, and smoking status). Green circles represent statistical significance at an unadjusted *P*<0.05.

