## Supplementary Materials for "Plasma proteome-wide analysis of cerebral small vessel disease identifies novel biomarkers and disease pathways"

### Supplementary Methods

***Selection of participants for ARIC MRI study***. 6,528 participants attended ARIC Visit 5. Of these participants, 1,978 received a 3T brain MRI. The participant selection criteria, which were described previously in Ducca 2021[1], are as follows. Participants with known MRI contraindications were excluded. MRI selection criteria included if they completed a brain MRI as part of the ARIC Brain MRI Ancillary Study in 2004-2006 or, demonstrated cognitive impairment. Cognitive impairment was defined as either a low Mini Mental Status Exam (MMSE) score (<21 for White participants and <19 for Black participants), or impairment on two or more cognitive domain scores at Visit 5 (<-1.5 standard deviations) and decline on the Delayed Word Recall test, Digit Symbol Substitution test, or Word Fluency test (Visit 5 score minus highest previous score <10th percentile on 1 or more tests or <20th percentile on 2 or more tests). An additional sample of cognitively intact participants with an age distribution that approximated that of the cognitively impaired participants were also selected.

***SomaScan protein quality control.*** The quality control steps performed by SomaLogic are detailed in a previous publication.[2] As part of the study-specific quality control, which is described elsewhere,[3] we ran blind duplicates for 187 of the 5,327 (4%) participants with available ARIC visit 5 SOMAmer measurements. The median inter-assay CV, calculated using the Bland-Altman method, was 4.7%. The median split sample reliability coefficient was 0.94 after excluding outliers. Of the 5,284 available SOMAmers, 94 were excluded for having a $CV_{BA}$ >50% or a variance of <0.01 on the log scale. In addition, 313 SOMAmers were excluded for binding to mouse Fc-fusion, a contaminant, or nonproteins. After QC, a total of 4,877 SOMAmers measuring 4,697 unique proteins or protein complexes were analyzed. Proteins were log2 transformed and outliers >5 SD away from the mean were winsorized. To establish the validity of SOMAmers binding to SVD-associated proteins, we used published SomaLogic validation data derived from multiple reaction monitoring mass spectrometry, data-dependent analysis mass spectrometry, Olink proximity extension assays, and identified cis pQTLs using GWAS (**Supplementary Table 21**). Twelve out of the 13 (92%) WMH-associated proteins were validated using one or more of these validation approaches.

***Ingenuity Pathway Analysis for SVD-associated proteins.*** To identify the biological pathways underlying the differential expression of SVD-associated proteins, we used Ingenuity Pathway Analysis (IPA). IPA is a bioinformatics platform that uses manually curated content from the Ingenuity Knowledge Base yielding insights from high-dimensional omics data (IPA, QIAGEN Inc., https://www.qiagenbioinformatics.com/products/ingenuity-pathway-analysis). We used the set of proteins associated with MRI-defined SVD variables at a two-sided $P$<0.01 for analyses. Beta coefficients (converted to log expression ratios) and $P$-values from the fully-adjusted model (model 2) were uploaded for each protein. Due to duplicate proteins mapping to the same gene, or more than one gene product corresponding to a single gene ID, not all examined proteins mapped to a unique gene in the IPA database. The gene ID was consolidated for proteins measured in duplicate, and the protein with the higher of the two expression values was used.

We performed IPA core analyses using the full set of genes in the Ingenuity Knowledge Base as a reference. We also repeated these analyses using the full set of SomaScan proteins as the reference set. Consistent with IPA parameters used previously, we considered both direct and indirect experimentally confirmed relationships from all species with a maximum of 35 molecules per network and 25 networks per analysis.[4,5] We performed a separate IPA canonical pathway analyses for the set of proteins associated with each MRI-defined SVD variable. One-sided *P*-values were calculated using a right-tailed Fisher's exact test to quantify the probability of overlap between the set of variable associated proteins and the protein set known to exist within a specific biological pathway. Statistical significance was set at an FDR-corrected $P<0.05$ threshold. Based on the directionality of the protein-specific associations with each outcome, a pathway-specific Z-score was calculated to quantify the likelihood that a particular pathway was activated or inhibited.

***Protein and gene expression in brain tissue:***

**Genotype-tissue expression analysis.** Using the GTEx project (version 8), a publicly available database of multi-tissue gene expression data[6], we assessed the expression of genes coding for WMH-associated proteins in whole blood, brain, and other tissue. GTEx protocols were approved by the NIH National Human Genome Research Institute; written informed consent was provided by all donors. We used the GTEx Multi Gene Query to visualize expression of WMH-associated protein-coding genes in terms of transcripts per million. Hierarchical cluster analysis was used to cluster genes and tissues based on expression.

**Protein-tissue expression analysis.** Using the Human Protein Atlas (version 20.1)[7], we assessed the expression of WMH-associated proteins in brain and other tissues. Specifically, the Tissue Atlas was used to provide information regarding the expression of human genes at the level of mRNA and protein across 48 normal human tissue types. Protein expression data, derived from donated tissue specimens, was quantified using antibody-based immunohistochemistry methods as either *low*, *medium*, *high*, or *not detected*. The human tissue samples used for protein expression analyses were acquired from the Department of Pathology, Uppsala University Hospital, Uppsala, Sweden as part of the sample collection governed by Uppsala Biobank (http://www.uppsalabiobank.uu.se/en/). Human tissue samples were collected in accordance with Swedish laws and regulation and were anonymized in accordance with approval and advisory report from the Uppsala Ethnical Review Board (Reference # 2002-577, 2005-338 and 2007-159 (protein) and # 2011-473 (RNA)). Donor information, including age and sex, can be located at http://www.proteinatlas.org/about/cellines.

**Brain cell-specific expression analysis.** Using Brain RNA-Seq (http://www.brainrnaseq.org/)[8], we assessed the expression of WMH-associated protein-coding genes at the mRNA level. Brain RNA-Seq—a database of human cell-type-specific gene expression data for astrocytes, neurons, oligodendrocytes, microglia/macrophage, and endothelial cells—used brain tissue samples obtained from patients undergoing neurosurgery to treat epilepsy or tumors. Tissue derived from normal temporal lobe cortex was resected to access deeper hippocampal regions. EEG and MRI confirmed the health of resected tissue. Mature human astrocytes, endothelial cells, microglia/macrophages, neurons, and oligodendrocytes were purified using protocol detailed previously.[8] Total RNA was extracted using the miRNeasy kit (QIAGEN); quality was determined using Bioanalyzer. RNA-seq reads were analyzed with the Galaxy web-platform (http://usegalaxy.org), and Cufflinks was used to assemble transcripts and estimate expression as

FPKM. Human brain tissue was obtained with informed consent under a protocol approved by the Stanford University Institutional Review Board.

***Identification of SVD-associated protein networks.*** The Netboost dimension-reduction procedure first uses the Spearman correlation coefficients from network protein-protein relationships to remove weak potentially spurious connections between proteins to reduce the network down to essential edges. This step reduces the noise and increases the robustness of the computed network and distance matrix. Second, a sparse version of hierarchical cluster and the Dynamic Tree Cut procedure was used to identify co-expressed protein subgroups (modules). Third, the first principal component score for the rank matrix of each protein module, referred to as the module eigenprotein (ME), was calculated as a value of person-specific protein module expression.[9,10] Consistent with previous proteomic network-based studies, we used a minimum module size of 20[11] and applied a Spearman filter method with a soft power of β=2, as defined by a network topology analysis that considered scale independence and mean connectivity. An "unsigned" network approach that does not consider the sign of the relatedness between two connected nodes was applied for clustering. The Netboost software used to create the protein modules is publicly available at (http://bioconductor.org/packages/release/bioc/html/netboost.html).

***Identification of instrumental variables (IVs) for WMH-associated proteins.*** To identify genetic loci for WMH-associated proteins and modules (protein quantitative trait locus (pQTL)), we performed genome-wide association analyses in the ARIC participants of European ancestry (N=3,583) using the 1000 Genome imputed genotype data and excluding SNPs with poor imputation quality (r2<0.5), significant Hardy-Weinberg disequilibrium test *P*-value ($<5x10^{-8}$) and low allele frequency (MAF<0.01). Genetic associations of each WMH-associated protein measured at ARIC visit 5 was estimated using PLINK v1.9[1]. Models were adjusted for sex, age and 3 genetic principal components (PCs). The genome-wide significant (*P*-value< $5x10^{-8}$) associations were then clumped at r2<0.05 using a web app, LDlink (11/06/2020 release)[2] against 1000 Genomes Project European reference panel to secure independence among instrument variables (IVs). Additional pQTL loci identified in the INTERVAL study[3] were also considered if they showed at least marginal association in ARIC (*P*-value<0.05). WMH IVs were identified using publicly available GWAS summary statistics published by Traylor et al. (2016).[5] We did not use the largest study (Murali et al.[6]) due to sample overlap with ARIC, but the identified loci were added as IVs if they were independent from other IVs and at least marginally significant in Traylor et al. When an IV was not included in the outcome data, we used a proxy (r2 > 0.8) within 500 kb.

***Functional annotation of pQTLs with deleteriousness and brain eQTL genes.*** We used the Functional Mapping and Annotation of Genome-Wide Association Studies (FUMA GWAS)[7] platform to annotate pQTLs that reached genome-wide significance. We examined whether pQTLs for WMH-associated proteins were also eQTLs for genes expressed in brain tissue using data from BRAINEAC[8], GTEx V83 brain tissue eQTL data[9], CMC[10], BrainSeq[11], and PSychENCODE[12]. Additionally, we examined whether the genes in brain tissue linked to the identified eQTLs (eGenes) were enriched using for biologic pathways and processes using MsigDB v7.0[13,14], GWAS catalog[15] and Wikipathways v20191010.[16]

***Variant Pathway Mapping.*** Variant-pathway mapping was used to assess the relationship between WMH-associated protein pQTLs and biological pathways of genes whose expression levels in the brain are associated with such genetic variation (eGenes). The aim of the approach was to capture information regarding biological pathways that might be affected by altered expression of WMH-associated protein-linked eGenes, thus identifying secondary biological correlates that may be associated with clinical outcomes. The current approach was developed based on its prior implementation[12]; however, it differs in that it is not restricted to a limited set of biological pathways selected *a priori*. The current approach computes standardized likelihoods between genetic variations (i.e., SNPs weighted according to their probability) and biological pathways (i.e., GO Terms weighted according to their probability) independent of the frequency of genes implicated in a given biological pathway, followed by correction for such frequencies to ensure reliability of results. This approach does not identify significantly enriched pathways as done in gene enrichment analysis, but rather captures the degree of association between genetic variation in each WMH-associated protein and the biological pathways of brain tissue eGenes. We first calculated a Weighted Variant-to-Gene Expression (VGE) Quotient [$\delta$] representing the probability of an association between a given brain eGene and genetic variation (SNPs) for a given WMH-associated protein (inferred from pQTLs). Specifically, we summed the frequency how many times a given brain eGene was associated with pQTLs of a given WMH associated protein. We then divided that frequency by the overall frequency of significant SNPs for a given WMH-associated protein. Next, we calculated a Variant-to-Pathway Estimate (VPE) [$\gamma$] representing the probability of an association between a WMH-associated protein pQTL and a biological pathway annotated for the corresponding eGene. Here, we multiplied the weighted probability of observing a given GO term for a given eGene by the Weighted Variant-to-Gene Expression (VGE) Quotient. Finally, we summed the VPE for each GO term for each WMH-associated protein to calculate a Variant-Pathway-Mapping (VPM) Total Probability, representing the degree of association between WMH-associated protein pQTLs and biological pathways linked to brain eGene expression. To ensure our VPM Total Probability was not dependent on the likelihood of a given biological pathway across our gene set, we corrected each VPM Total Probability value (i.e., VPM(Adjusted)) by dividing it by the frequency of genes annotated for a given biological pathway.

***External Replication:***

**Baltimore Longitudinal Study of Aging (BLSA).** The BLSA is an ongoing longitudinal study of community-dwelling volunteers conducted in Baltimore, Maryland USA. The BLSA cohort, participant enrollment criteria, and study procedures have been described in detail previously.[13] BLSA has a continuous enrollment; therefore, participants entered the study at different times. All BLSA participants began receiving repeat 3T MRIs as early 2009. Participant visits, which include a clinical exam, a cognitive exam, and brain imaging (including MRI), take place every 1 to 4 years according to participant age (age <60, every 4 years; age 60-79, every 2 years; age ≥80, every year). The BLSA protocol has been approved by the Institutional Review Board of the National Institute of Environmental Health Science, National Institutes of Health (03AG0325). Informed consent was obtained from all participants. Deidentified data were used for all BLSA analyses.

Proteins were measured in plasma collected at the time of the earliest available 3T MRI visit using a standardized protocol. Proteins were measured using the SomaScan platform (version 4.1). BLSA plasma specimens were shipped on dry ice to SomaLogic for quantification. Of the 1704 unique samples provided for proteomic measurement, 7 were excluded for failing to pass SomaLogic's quality control criteria. The 13 candidate proteins examined in BLSA had an intra-assay CV <8% as defined using a set of 102 blind duplicates. Log2 transformation was applied to all protein measurements to correct for skewness. For each protein, we winsorized outliers that were greater or less than 5xSD from the sample mean on the log2 scale. A 3T Achieva MRI was used to obtain T1-wieghted magnetization-prepared rapid gradient echo (MPRAGE) scans. The Multi-atlas Region Segmentation Utilizing Ensembles (MUSE) anatomic labeling method was used to generate an ensemble of labeled atlases in target image space.[14] This approach, which has been defined in detail elsewhere, was used to define the total volume of white matter. WMH volume was defined by FLAIR scans using DeepMRSeg to segment white matter lesions.[15] WMH volume was log transformed to correct for skewness.

We used separate multivariable linear regression models to relate log2 protein levels to MRI measures of total WMH volume and total white matter volume. Models were adjusted for intracranial volume, age, sex, race, education, *APOE*ε4 genotype, eGFR, and a comorbidity score. The comorbidity score was calculated based on the presence/absence of eight chronic diseases at the time of brain MRI (i.e., hypertension, diabetes mellitus, obesity, ischemic heart disease, congestive heart failure, cancer, chronic obstructive pulmonary disease [COPD], and chronic kidney disease [CKD]).

**Generation Scotland (GS).** Study participants were recruited for the 'STratifying Resilience and Depression Longitudinally' (STRADL) study from the Generation Scotland: Scottish Family Health Study (GS). The GS study is a large, population-based cohort study of over 24,000 individuals residing in Scotland. Recruitment was completed between 2006 and 2011; a clinical visit during this time collected detailed health and cognitive tests as well as biological samples, including blood, urine, saliva. Complete details of the STRADL cohort and GS protocol have been published previously.[16–18] Briefly, blood was collected into standard polypropylene EDTA test tubes; following centrifugation, plasma was aliquoted in 500 μL aliquots and stored at −20°C for future analyses.[18] Written informed consent was provided by all participants; ethical approval for STRADL was formally obtained from the NHS Tayside committee.

A total of 1061 cognitively normal individuals (59% female) were selected from STRADL study. Plasma data, demographic information, and cognitive tests (digit symbol, verbal fluency, Mill Hill vocabulary scale and Wechsler memory test) were administered to all subjects.[16] Fluid-attenuated inversion recovery (FLAIR) scans, available for 940 subjects, were utilized to obtain a Fazekas score, an index of white matter hyperintensities.

Plasma proteins were measured using the SOMAscan platform (version 4.0).[19] After raw data processing and quality control measures, 4235 proteins were available for analysis. Each protein was log-transformed; effects of sample collection site and plasma storage time on proteins were accounted for by linear regression and the residuals were incorporated into subsequent analyses.

WMHs were identified using FLAIR scan files and were defined as punctuate, focal or diffuse lesions in the deep or periventricular white matter, basal ganglia or brainstem, visible as areas of hyperintensity on FLAIR images. Severity of WMHs was graded according to the Fazekas scale,[20] which grades periventricular WMHs and deep WMHs from 0 (absent) to 3 (severe). A score of 1 is defined as caps or pencil thin lining (periventricular WMHs) or punctuate foci (deep WMHs); a score of 2 is defined as a smooth halo (periventricular WMHs) or beginning to confluence (deep WMHs) and a score of 3 is defined as irregular periventricular signal extending into the deep white matter (periventricular WMHs) or large confluent areas (deep WMHs). In the current analyses, total scores—calculated by summing the periventricular and deep white matter scores—were used. Partial Spearman correlation tests were used to evaluate the association between proteins and WMHs (Fazekas scores), adjusting for age, sex and *APOE* ε4 genotype. *P* values were corrected using a false discovery rate (FDR) for multiple testing.

**Cardiovascular Health Study (CHS).** The Cardiovascular Health Study (CHS) is a population-based cohort study of risk factors for coronary heart disease and stroke in adults ≥65 years conducted across four communities.[21] In June 1990, four Field Centers (Sacramento, CA; Hagerstown, MD; Winston-Salem, NC; Pittsburgh, PA) completed the recruitment of 5201 primarily European ancestry participants from random sampling of Medicare eligibility lists. Between November 1992 and June 1993, an additional 687 African Americans were recruited using similar methods for a total sample of 5,888. Blood samples were drawn from all participants at their baseline examination and during follow-up clinic visits. Between enrollment and 1998-99, participants were seen in the clinic annually, and contacted by phone at 6-month intervals to collect information about hospitalizations and potential cardiovascular events. Participants with previously unthawed plasma blood samples from the 1992-3 clinic visit were included for proteomics analysis. CHS was approved by institutional review committees at each field center and individuals in the present analysis gave informed consent including consent for the study of cardiovascular disease.

Proteomics assays were performed at SomaLogic among participants who consented and who had previously unthawed plasma blood samples using the SomaScan platform (version 4.0) (n = 3656).[22] *CHS brain imaging.* Two brain MRI scans were performed: the first scan was completed in 3,660 participants in CHS years 4-6, and the second scan was completed in 2,317 participants in years 10-11. The median and mean time between initial and follow-up scans was 5 years; 2,116 participants received both scans. Both MRI scans included sagittal $T_1$-weighted localizer sequences, and axial $T_1$, spin-density, and $T_2$-weighted images.[23,24] Neuroradiologists, blinded to the subjects' age, sex, race, ethnicity, and clinical information, evaluated the images using a standardized protocol.[23,24] White matter grade (WMG) was assessed by comparing the burden of periventricular and subcortical white matter-signal abnormality on either axial $T_2$- weighted or spin-density images to a series of eight images with successively increased white matter changes from barely detectable to extensive.[23] WMH was graded using a 10-point semiquantitative scale from a white matter grade (WMG) of 0 (least severe) to 9 (most severe). For WMG, there was an interreader intraclass correlation coefficient of 0.76 and an intrareader coefficient of 0.89.[23] To determine WMG progression, scans were re-read side-by-side by neuroradiologists who were blinded to previous grade determination and order of scans; these reads demonstrated an intrareader reliability kappa of 0.59, and inter-reader reliability kappa of 0.36; certain scans were reviewed and adjudicated, as previously discussed.[24] WMH volume was derived for a subset of participants who received imaging on 1.5T scanners, which included 3D T1-weighted spoiled gradient-recalled (SPGR) sequences, at the follow-up MRI visit (10-11 years). FreeSurfer software was used to derive WMH volume from T1-weighted sequences. Of the 765 participants with proteomic data who completed a follow up MRI and had complete covariate information, 680 had an initial MRI available to assess worsening WMG.

We used multivariable linear regression models to relate standardized log2 protein levels to MRI measures of total WMH volume. Multivariate poisson regression was used to assess risk of worse WMG on the follow up MRI compared to the first MRI. Models were adjusted for age, sex, race, education, and eGFR, BMI, diabetes, hypertension, and smoking status at the time of protein measurement. WMH volume was log transformed and analysis also included adjustment for intracranial volume and time between blood draw for proteomic assessment and MRI. Worsening WMG included adjustment for time between the first and follow up MRI.
